# The role of sleep traits in prostate, endometrial, and epithelial ovarian cancers: An observational and Mendelian randomisation study

**DOI:** 10.1101/2025.04.10.25325598

**Authors:** Christos V. Chalitsios, Eirini Pagkalidou, Christos K. Papagiannopoulos, Georgios Markozannes, Emmanouil Bouras, Eleanor L Watts, The Practical Consortium, Rebecca C. Richmond, Konstantinos K. Tsilidis

## Abstract

**Background:** Sleep traits may influence cancer risk; however, their associations with prostate (PCa), endometrial (ECa), and epithelial ovarian (EOCa) cancer remain unclear.

**Methods:** We conducted an observational analysis using the UK Biobank cohort and a two-sample Mendelian randomisation (MR) analysis to investigate the association of six sleep traits-duration, chronotype, insomnia, daytime napping, daytime sleepiness, and snoring-with PCa, ECa, and EOCa risk. Cox proportional hazards models were used for the observational analysis, while the inverse variance-weighted (IVW) method was applied in MR, with multiple sensitivity analyses. A Bonferroni correction accounted for multiple testing.

**Results:** Among 8,608 PCa, 1,079 ECa, and 680 EOCa incident diagnoses (median follow-up: 6.9 years), snoring was associated with reduced EOCa risk (HR=0.78, 95%CI: 0.62–0.98), while daytime sleepiness was associated with increased EOCa risk (HR=1.23, 95%CI: 1.03-1.47). However, these associations were not confirmed in MR. MR suggested higher odds of PCa (OR_IVW_=1.05, 95%CI: 1.01-1.11) and aggressive PCa (OR_IVW_=1.10, 95%CI: 1.02-1.19) for evening compared to morning chronotype. None of the findings survived multiple testing correction.

**Conclusion:** Sleep traits were not associated with PCa, ECa, or EOCa risk, but evening chronotype may increase PCa risk. Further research is needed to verify this association and investigate potential underlying mechanisms.

**Impact:** The proposed results have potential utility in reproductive cancer prevention.

**What is already known on this topic:** Sleep traits have been implicated in cancer risk, but their associations with prostate, endometrial, and epithelial ovarian cancer remain unclear.

**What this study adds:** This study found suggestive evidence that an evening chronotype may be associated with an increased risk of overall and aggressive prostate cancer.

**How this study might affect research, practice or policy:** Further research is needed to confirm the potential association between chronotype and prostate cancer risk, which could inform personalised cancer prevention strategies.

## INTRODUCTION

Sleep is a complex, reversible neurobiological state marked by distinct brain and body activity patterns, leading to temporary disengagement from the environment (1). Essential for normal physiology, it influences growth hormone secretion (2), physical repair (3), immune function (4,5), and metabolism (3). Sleep is regulated by homeostatic and circadian processes (6) and includes dimensions such as duration, quality (e.g., insomnia, snoring), and chronotype (7). Single-nucleotide polymorphism (SNP)-based studies estimate self-reported sleep traits have 5–15% heritability (8–11).

Healthy sleep entails sufficient duration, good quality, appropriate timing, regularity, and absence of disturbances (12). Nevertheless, up to 70 million people in the U.S. and 45 million people in Europe suffer from chronic sleep disorders (13,14). Abnormal sleep patterns—insufficient duration, poor quality, or irregular timing—are associated with adverse health outcomes (15), including breast (16) and lung (17,18) cancers. Potential mechanisms involve circadian rhythm disruption (19–21), neuroendocrine and immune pathway alterations (19,20), and cancer-stimulatory cytokines (22).

Despite sleep’s role in health, its link to reproductive cancers remains unclear. A meta-analysis found no association between insomnia and prostate cancer (PCa) (2 studies, 4,909 cases), while a UK cohort (23) with more incident cases (n=6,747) reported a higher PCa risk (23,24). Mendelian randomisation (MR) studies found that the morning preference was associated with a lower PCa risk (25,26), but cohort studies could not establish a link (23,27,28). Research on endometrial (ECa) and epithelial ovarian cancer (EOCa) is limited. No study has examined insomnia or snoring and ECa, while one MR study found an association between insomnia and increased endometrioid EOCa risk but decreased high-grade serous and clear cell EOCa risk (29).

Given the inconsistent and limited evidence currently available, further research is needed. We examined six sleep traits -sleep duration, chronotype, insomnia, daytime napping, daytime sleepiness, and snoring- and their association with PCa, ECa, and EOCa. Our study included an observational analysis in the UK Biobank (UKB) and a two-sample MR study using sex-combined and sex-specific estimates based on the largest genome-wide association studies (GWAS).

## MATERIALS and METHODS

### Observational analysis

#### Data source and study population

We used data from all UK Biobank (UKB) participants (30), a prospective cohort of over 500,000 participants aged 37-73 years, recruited from 22 UK centres between 2006 and 2010. We excluded individuals of non-white ancestry, those with aneuploidy (putative sex chromosome configurations other than XX or XY), and participants with mismatched self-reported and biological sex (Figure S1). Women with a hysterectomy or oophorectomy were also excluded. To minimise reverse causation, we excluded participants with cancer at baseline or rare cancer histology. Lastly, individuals with extreme body mass index (BMI) (±6 SD from the mean) were removed.

#### Sleep traits

At baseline, participants completed a touchscreen questionnaire on sleep duration, chronotype, insomnia, daytime napping, daytime sleepiness, and snoring.

Sleep duration was assessed by asking: “*About how many hours of sleep do you get every 24 hours? (please include naps)*”. We examined sleep duration continuously and categorically (short [<7 hours/day], normal [7-8 hours/day], and long [>8 hours/day]).

Chronotype was assessed in the question “*Do you consider yourself to be*?” with one of six possible answers: “*Definitely a ‘morning’ person*”, “*More a ‘morning’ than ‘evening’ person*”, “*More an ‘evening’ than a ‘morning’ person*”; “*Definitely an ‘evening’ person*”, “*Do not know*”, or “*Prefer not to answer*”. In addition to the above categories, we dichotomised chronotype into morning *(“definitely a ‘morning’ person”* or *“more a ‘morning’ than ‘evening’ person”*) or evening *(“definitely an ‘evening’ person”* or *“more an ‘evening’ than ‘morning’ person”*) preference.

To assess insomnia, participants were asked: “*Do you have trouble falling asleep at night or do you wake up in the middle of the night?*” with responses “*Never/rarely*”, “*Sometimes*”, “*Usually*”, or “*Prefer not to answer*”. In addition to the above categories, we dichotomised insomnia into yes (“*sometimes”* or “*usually*”) and no (“*never/rarely*”).

Daytime napping and daytime sleepiness were defined based on the questions *"Do you have a nap during the day?"* and *"How likely are you to doze off or fall asleep during the daytime when you don’t mean to? (e.g. when working, reading or driving)",* with responses “*Never/rarely*”, “*Sometimes*”, “*Usually*”, or “*Prefer not to answer*”. In addition to the above categorisation, responses were coded continuously, corresponding to severity.

Snoring was assessed with the question: “*Does your partner or a close relative or friend complain about your snoring*?” with responses “*Yes*”, “*No*”, “*Do not know*”, or “*Prefer not to answer*”.

Participants reporting “*do not know*” and “*prefer not to answer*” were set to missing, unless otherwise specified.

#### Covariates

Through interviews and questionnaires, information was collected at baseline, covering various aspects, including demographics (age, sex), socio-economic (Townsend deprivation index, Education), lifestyle characteristics (smoking status, coffee intake, tea intake, metabolic equivalent of task [MET]), anthropometric measures (BMI), family history of cancer, and sex-specific factors (female: menopausal status, hormone replacement therapy [HRT], parity; male: history of prostate-specific antigen [PSA] testing) (Table S1). Sleep apnoea was ascertained using the International Classification of Diseases 10th (ICD-10) revision codes (G47.3) and record-linkage data from local general practitioners.

#### Reproductive system cancers

Cancer incidence was obtained via linkage to national registries in England, Wales, and Scotland. The ICD-9 and ICD-10 codes were employed to define PCa, ECa, and EOCa. Malignant cancers were classified using behavior codes 3 (malignant, primary site) or 5 (malignant, microinvasive) and ICD-9/ICD-10 codes defined prostate (C61/185), endometrial (C54/182), and ovarian (C56/183) cancers. First primary incident cases were identified using diagnosis date, ICD-9/10 codes, morphology, and histology (31). To avoid misclassification, diagnoses were included until December 31, 2020, to ensure data completeness (Figure S2).

#### Statistical analysis

Multivariable Cox proportional hazards models estimated hazard ratios (HRs) for associations between sleep traits and PCa, ECa, and EOCa risk. Schoenfeld residuals tested proportional hazards assumptions. Follow-up was from recruitment to cancer diagnosis, death, or last follow-up (December 31, 2020).

Initial models adjusted for age, BMI, and Townsend index. Fully adjusted models included additionally smoking, coffee/tea intake, education, and physical activity. For ECa and EOCa, sex-specific adjustments were made for menopause status and hormone replacement therapy (HRT), while for PCa, adjustment was made for prostate-specific antigen (PSA) testing. Snoring models adjusted for sleep duration, insomnia, and sleep apnea in sensitivity analyses.

Subgroup analyses were based on median age at recruitment and BMI. Non-linear associations between sleep duration and PCa, ECa, and EOCa risk were explored with Cox models using restricted cubic splines (knots at 5th, 50^th^ [reference], and 95th percentiles).

Three sensitivity analyses ensured robustness: (1) excluding participants with less than two years of follow-up to reduce reverse causation, (2) excluding those with diagnosed sleep disorders (ICD-10: G47) or night-shift work history, and (3) fully adjusted models also included alcohol intake as a covariate.

To account for multiple comparisons, a Bonferroni-corrected significance threshold of 0.003 (0.05/6[exposures)]x3[outcomes]) was applied. Results with p-values between this threshold and the nominal significance were considered suggestive. The data were managed and analysed using R Statistical Software (v4.4.0; R Core Team 2024) (32).

### Mendelian randomisation study

#### Sleep trait GWAS

Sleep duration was assessed at baseline assessment in UKB and was treated as a continuous variable. Self-reported short sleep (<7 hours vs. 7-8 hours; *n* = 106,192 cases and 305,742 controls) and long sleep duration (>8 hours vs. 7-8 hours; *n* = 34,184 cases and 305,742 controls) were also evaluated in participants of European ancestry from UKB (8). Dashti *et al.* (8) performed a GWAS among UKB participants only (*n*=446,118).

Chronotype was assessed at baseline in UKB as described above and in 23andMe (33) via a single question, “*Are you naturally a night person or a morning person?*”. To maximise statistical power, Jones *et al.* (10) dichotomised chronotype into morning or evening preference and used results from both UKB and 23andMe in a GWAS meta-analysis (n=697,828).

Insomnia was assessed at baseline in UKB, as described in the observational analysis section above. Participants in 23andMe (33) were asked to answer one or more questions about seven sleep-related traits. Participants with a positive response to any of the following questions were considered as cases: 1) ‘*Have you ever been diagnosed with, or treated for, insomnia?*’, 2) ‘*Were you diagnosed with insomnia?*’, 3) ‘*Have you ever been diagnosed by a doctor with any of the following neurological conditions?*’, 4) ‘*Do you routinely have trouble getting to sleep at night?*’, 5) ‘*What sleep disorders have you been diagnosed with? Please select all that apply. (Insomnia, trouble falling or staying asleep)’*, 6) ‘*Have you ever taken sleep aids medications?’* and 7) ‘*In the last two years, have you taken any sleep aids medications?*’’. To maximise statistical power, Watanabe et al. (11) dichotomised insomnia status (yes/no) and conducted a GWAS meta-analysis combining data from both the UK Biobank and 23andMe cohorts (n 2,365,010).

Daytime napping and sleepiness were assessed in UKB, as described above. GWAS analyses by Dashti et al. (34) and Wang et al. (35) included sample sizes of 452,633 and 452,071, respectively. Responses were coded continuously based on severity.

Snoring was assessed at recruitment in UKB as above, and Campos *et al.* (11) performed a GWAS (n=408,317).

#### Genetic variant selection

Instrumental variables (SNPs) were selected based on genome-wide significance (P<5×10⁻D) to represent genetic susceptibility to sleep traits. Variants unavailable in outcome datasets, palindromic SNPs, those in linkage disequilibrium (LD) (r²≥0.001, ±10,000 kb), or with an F-statistic<10 were excluded (36). Primary analyses used sex-specific GWAS instruments, with secondary analyses using sex-combined instruments to enhance statistical power at the cost of sex-specific precision.

#### Reproductive system cancers GWAS

Summary statistics for ECa risk were obtained from the Endometrial Cancer Association Consortium (ECAC) (12,906 cases) (37). Subtype-specific data included endometrioid (8,758 cases) and non-endometrioid (1,230 cases) histologies, confirmed via pathology reports (37).

PCa GWAS summary statistics were derived from the PRACTICAL Consortium (38), the largest GWAS meta-analysis on PCa (79,148 cases of European ancestry). We also obtained summary statistics on aggressive PCa (15,167 cases), defined as metastatic disease, a Gleason score≥8, a PSA>100 ng/mL, or PCa-related death.

EOCa genetic data were sourced from the Ovarian Cancer Association Consortium (OCAC) (39), including 25,509 cases (22,406 invasive and 3,103 low malignant potential). Subtype-specific data were available for serous EOCa (16,003 cases).

#### Statistical analysis

The primary two-sample MR method was the random-effects inverse variance–weighted (IVW) model, with three sensitivity analyses (weighted median (40), weighted mode (41), MR-Egger (42)) (Figure S3). The MR pleiotropy residual sum and outlier test (MR-PRESSO) was used to detect and exclude outlier SNPs by applying a random-effects IVW model (43). To assess potential non-linear associations of sleep duration with ECa, PCa, and EOCa risk, two-sample MR analyses used genome-wide significant SNPs for short sleep (<7 vs. 7–8 hours) and long sleep (>8 vs. 7–8 hours).

A Bonferroni correction set statistical significance at P<0.0042 (0.05/4 exposures × 3 outcomes), with results between this and nominal significance considered suggestive. MR analysis was performed with R Statistical Software (v4.4.0; R Core Team 2024) (32) using the “TwoSampleMR” package.

## RESULTS

### Observational analysis

After applying exclusion criteria in UKB, 8,608 of 196,385 white men developed incident PCa, 1,079 of 183,662 white women developed incident ECa, and 680 of 183,662 white women developed incident EOCa over median follow-ups of 6.9, 6.2, and 6.4 years, respectively (Table 1).

**Table 1.**
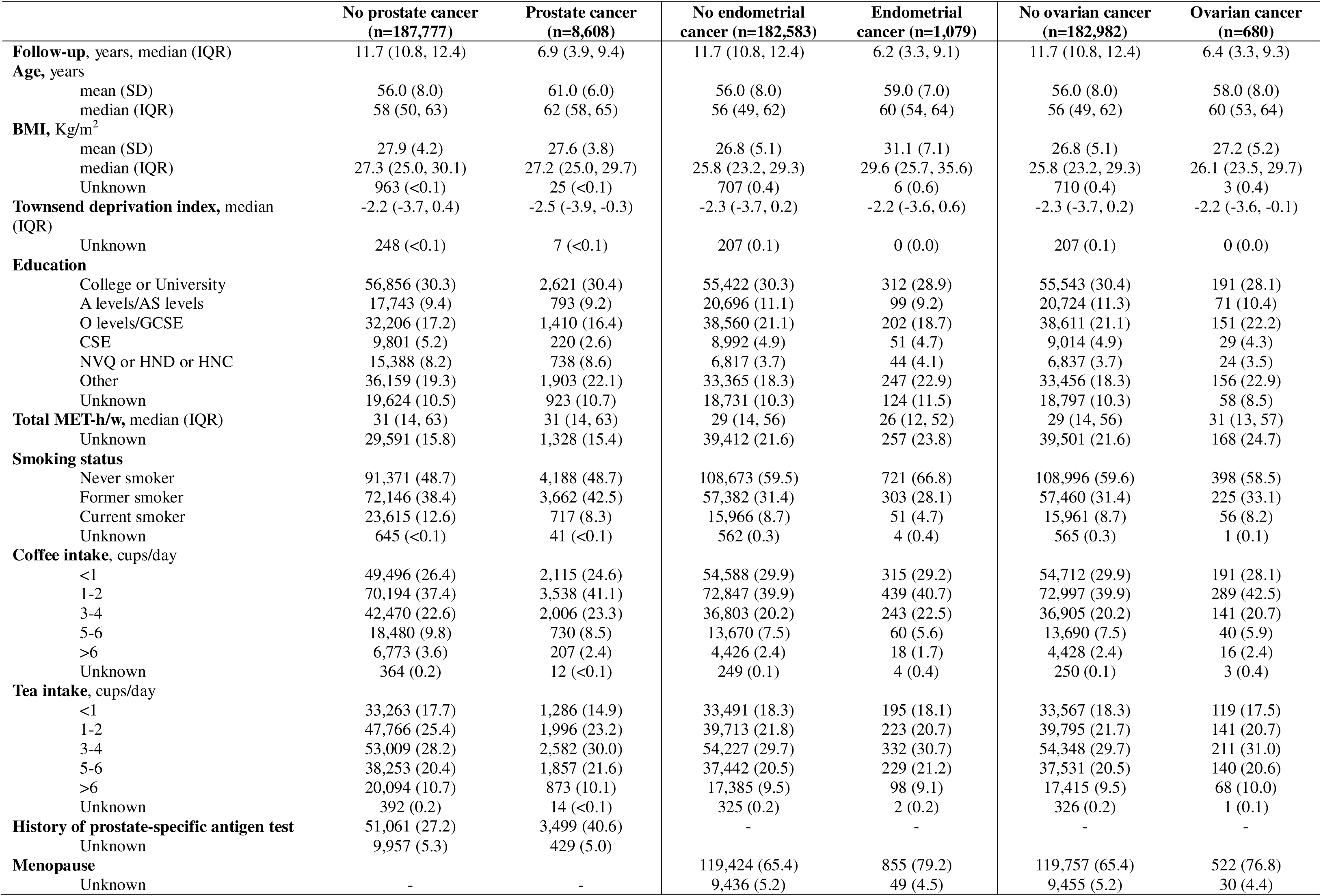
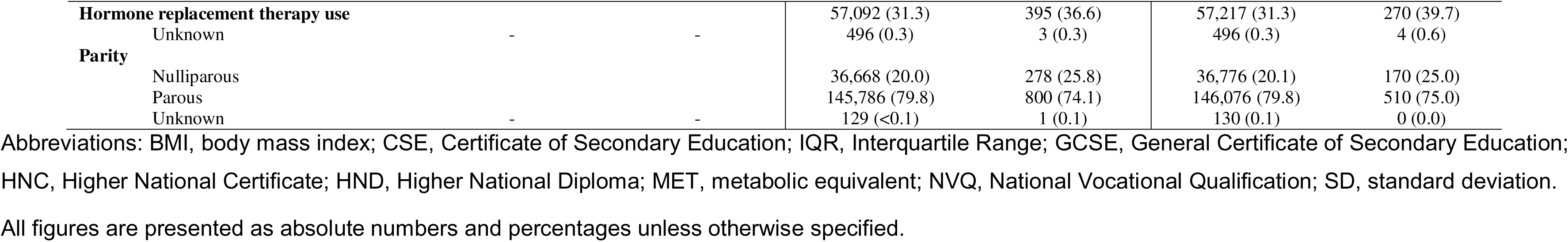
Baseline characteristics of study participants stratified by type of reproductive system cancer in the UK Biobank.

Individuals with cancer were generally older than those without (Table 1). Mean BMI was higher in women with ECa than without (31.1 kg/m² vs. 26.8 kg/m²). No major differences were observed in education, physical activity, or Townsend deprivation index between cancer and non-cancer groups. PSA testing was more common among those with PCa (40.6% vs. 27.2%). More women with ECa or EOCa were postmenopausal compared to those without these cancers. Baseline characteristics stratified by sleep traits are detailed in Tables S2–S4.

Sleep duration, daytime napping, and insomnia showed no association with the risk of PCa, ECa, and EOCa (Figure 1, Table S5). Snoring was associated with lower EOCa risk (HR=0.78, 95%CI: 0.62-0.98), while daytime sleepiness was associated with higher EOCa risk (HR=1.23, 95%CI: 1.03-1.47). Chronotype was not associated with cancer risk, but in younger participants (≤58 years), an evening chronotype had a 14% higher PCa risk (HR=1.14, 95%CI: 1.03-1.26) compared to a morning chronotype (P_interaction(≤58 vs. >58 yrs old)_=0.046) (Table S6). No other notable subgroup differences were observed (Table S7-S11). No evidence of a non-linear relationship between sleep duration and cancer risk was found (Figure S4) and results remained consistent in sensitivity analyses (Tables S12 & S13). However, all the above findings did not withstand correction for multiple comparisons.

**Figure 1.**
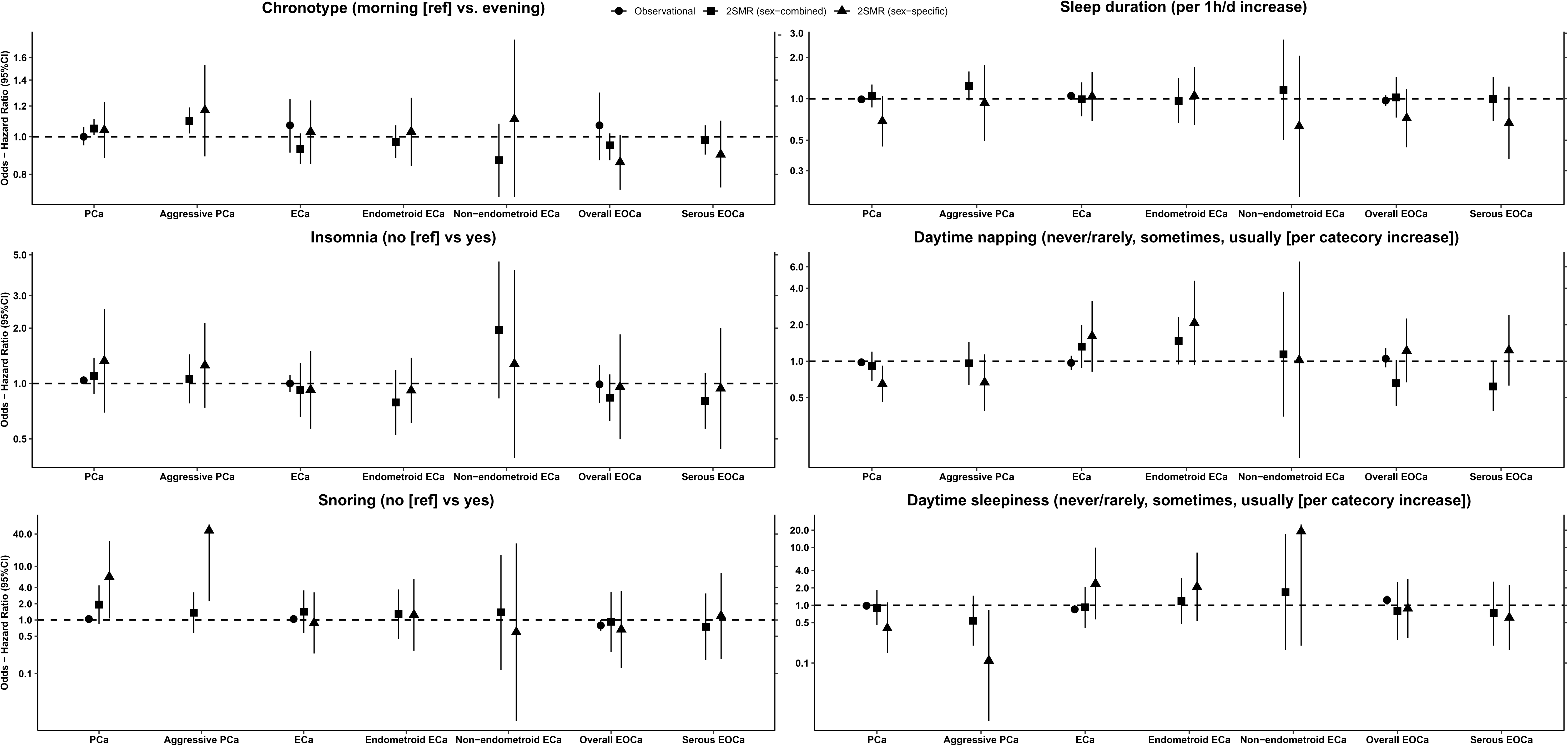
Forest plots illustrate the associations between sleep traits and the risk of prostate cancer (PCa), endometrial cancer (ECa), and epithelial ovarian cancer (EOCa). Hazard ratios (HR) and 95% confidence intervals (CI) were estimated using Cox proportional hazard models adjusted for age, body mass index (BMI), Townsend deprivation index, smoking status, coffee intake, tea intake, education level, and physical activity (MET-hours/week). Additional sex-specific adjustments were made for menopause status and hormone replacement therapy (HRT) in ECa and EOCa analyses and for prostate-specific antigen (PSA) testing in PCa analyses. Odds ratios (OR) and 95% CI were estimated using a two-sample Mendelian randomisation (MR) analysis (IVW method) to assess relationships between genetically proxied sleep traits and cancer risk.

### Mendelian randomisation

Consistent with the observational analysis, MR found no evidence associating sleep duration, insomnia, daytime napping, or daytime sleepiness with cancer risk (Figure 1, Table S14). However, genetic predisposition to an evening chronotype was associated with higher odds of PCa (OR_IVW_=1.05, 95%CI: 1.01-1.11) and aggressive PCa (OR_IVW_=1.10, 95%CI: 1.02-1.19) compared to a morning chronotype, with directionally consistent results in sensitivity analyses (Table S14). In contrast to observational findings, genetically predicted snoring was not associated with EOCa (OR_IVW_=0.92, 95%CI: 0.26-3.36). However, none of these associations remained significant after multiple comparison correction. Sex-specific analyses aligned with the main findings but were less precise (Figure 1, Table S15).

## DISCUSSION

We investigated the association between six sleep traits and the risk of PCa, ECa, and EOCa using data from UKB. To triangulate evidence, we performed MR using genetic variants associated with these sleep traits from large-scale GWAS. Our findings suggest a genetically predicted evening chronotype may be associated with a higher risk of overall and aggressive PCa. In the observational analysis, younger participants (≤58 years) with an evening chronotype had a higher PCa risk. This was supported by Mendelian randomisation, where evening chronotype was found to increase risk of overall and aggressive cancer (more common in younger men). Snoring and daytime sleepiness were associated with EOCa in observational analyses, but MR did not support these findings. None of the associations survived correction for multiple comparisons.

Several studies (23,27,44–51) have explored sleep duration, insomnia, snoring, daytime napping and sleepiness in prostate carcinogenesis without finding any association, consistent with our results. Previous observational studies (23,27,28) including ours, found no association between chronotype and PCa risk. However, our MR analysis suggested a lower PCa risk for morning chronotype, consistent with two prior MR studies (25,26), and we additionally observed a lower aggressive PCa risk. Genes near chronotype-related variants may contribute to PCa pathophysiology. Specifically, the *LRPPRC* gene, encoding a leucine-rich protein, may inhibit apoptosis in PCa cells (52), while *MEIS1* depletion has been associated with increased prostate tumor growth (53,54). A plausible explanation for these associations is that evening chronotype may exacerbate circadian disruption, increasing PCa risk (55). A prior MR examined testosterone as a mediator, but findings remain inconclusive (26). Further research is needed to clarify potential mechanisms.

In line with the Women’s Health Initiative (WHI) Observational Study (56), a pooled analysis of US and European studies (57), and two Japanese cohorts (58), we found no association between sleep duration and ECa risk, whether analysed continuously or categorically. Notably, the pooled analysis (57) suggested an inverse association in obese women (BMI≥30 kg/m²), but this was not confirmed in our study or WHI (56), although the direction was consistent. Discrepancies may reflect limited power in our study (159 cases) and WHI (212 cases) compared to the pooled analysis (2,089 cases). A potential mechanism is that extended sleep may increase melatonin (59), counteracting the higher circulating estrone levels in obese postmenopausal women (60). We found no association between insomnia, snoring, and ECa in observational or MR analyses, but prior studies on these relationships are scarce. Our findings also do not support a role for chronotype in ECa carcinogenesis, aligning with previous research (28,61).

Consistent with previous studies (28,44,45,62,63), we found no association between sleep duration or chronotype and EOCa. A Chinese cohort (44) found no association between insomnia and EOCa, supporting our results, while a Korean cohort (64) reported a borderline inverse association. MR analysis also showed no association between insomnia and overall or serous EOCa. Interestingly, another MR study (29) suggested insomnia increased endometrioid EOCa risk but decreased high-grade serous and clear cell EOCa risk, possibly indicating subtype-specific effects. These variations in findings across studies might reflect differences in the impact of insomnia on EOCa subtype. In our observational analysis, snoring was inversely associated with EOCa risk, but this was not supported by MR and did not survive multiple testing correction. The only other study on this topic (44) found no association. Given the inconsistent evidence and unclear biological plausibility, further research is needed.

A key strength of this study is the large sample size and multidimensional approach, integrating observational and MR analyses. Examining multiple sleep traits allowed for a comprehensive evaluation of sleep and reproductive cancer risks. MR helped address limitations of observational studies, such as reverse causation and residual confounding, assuming its core assumptions were met (65,66). However, several limitations should be considered. UKB self-reported sleep traits may be subject to measurement and recall bias. For instance, sleep duration may capture napping, while insomnia was assessed via reported symptoms, which may not fully align with clinical definitions. Although we adjusted for multiple confounders, residual confounding cannot be ruled out. Sensitivity analyses did not alter results, strengthening their robustness. While we tested MR assumptions via sensitivity analyses, we cannot be certain they were fully met. Using summary data limited our ability to assess non-linear sleep duration– cancer relationships. One-sample MR was not feasible due to limited power, but we accounted for non-linearity by using instruments for short and long sleep. Finally, as the UK Biobank does not represent the general population and predominantly includes European participants aged 38 to 73, caution is needed when generalising these findings to other age groups or populations.

In conclusion, we found no associations between six sleep traits and the risk of PCa, ECa and EOCa. However, a suggestive association emerged between evening chronotype and a higher risk of overall and aggressive PCa. Future research should replicate this association and uncover potential underlying mechanisms.

## Data availability

This observational analysis study was conducted under the UK Biobank application number 79696. Access to the data used in this study is subject to approval by UK Biobank and requires an application process. The data are not publicly available but can be accessed by bona fide researchers through the UK Biobank Access Management System (www.ukbiobank.ac.uk).

## Author Contributions

C.V.C and E.P. had full access to all the data in the study and are responsible for the integrity of the data and the accuracy of the data analysis.

Concept and design: C.V.C, G.M, R.C.R, K.T.T.

Acquisition of the data: C.V.C, C.P.

Statistical analysis: C.V.C, E.P.

Drafting of the manuscript: C.V.C.

Critical revision of the manuscript: All authors.

Acquisition of the financial support for the project leading to this publication: C.V.C.

## Acknowledgements

The authors thank the World Cancer Research Fund (WCRF) for funding this research, the staff and participants who contributed to the UK Biobank study, and the authors of the cited genome-wide association studies for sharing the summary statistics data. Acknowledgement for the PRACTICAL Consortium is provided in the Supplements.

## Ethics statement

The UK Biobank obtained ethical approval from the North West Multi-Centre Research Ethics Committee (approval number: 11/NW/0382) and acquired informed consent from all participants. This observational analysis study was conducted under the UK Biobank application number 79696. The Mendelian randomisation analyses do not involve human subjects, any anonymised individual patient data, or interaction/intervention with human subjects. This study used summary data published by multiple GWAS; patient consent was obtained by the corresponding GWAS. Consequently, there was no need for ethical approval.

