## Supplements for "The role of sleep traits in prostate, endometrial, and epithelial ovarian cancers: An observational and Mendelian randomisation study"

**Supplementary material**

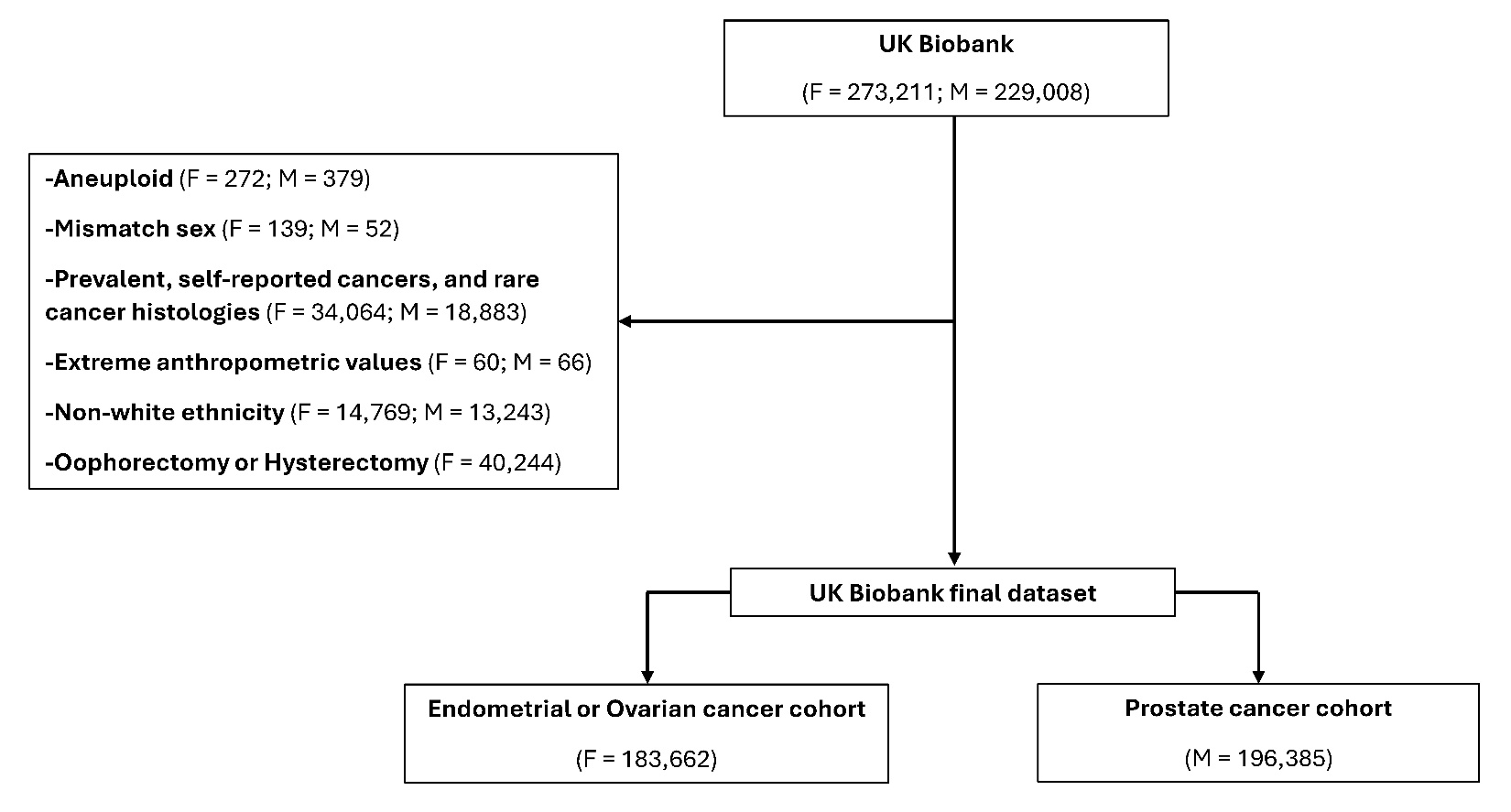

**Figure S1**. Flow chart of the observational analysis.

**Table S1**. Covariate and exposure variables used in the cohort study as they defined in the UK Biobank.

| **Category** | **Variable name** | **Data field** | **Description** |
| --- | --- | --- | --- |
| Covariate | Age | 21022 | Age at recruitment |
| Covariate | Body mass index | 21001 | Body mass index |
| Covariate | Smoking status | 20116 | Smoking status |
| Covariate | Alcohol intake | 1558 | Alcohol intake frequency |
| Covariate | Townsend deprivation index | 22189 | Townsend deprivation index at recruitment |
| Covariate | Coffee intake | 1498 | Coffee intake |
| Covariate | Tea intake | 1488 | Tea intake |
| Covariate | Total MET-h/w | 22038 | MET minutes per week for moderate activity |
|  |  | 22039 | METminutes per week for vigorous activity |
|  |  | 22037 | MET minutes per week for walking |
| Covariate | Education | 6138 | Qualifications |
| Covariate | Prostate-specific antigen test history | 2365 | Ever had prostate specific antigen (PSA) test |
| Covariate | Age at menarche | 2714 | Age when periods started (menarche) |
| Covariate | Oral contraceptive use | 2784 | Ever taken oral contraceptive pill |
| Covariate | Menopause | 3581 | Age at menopause (last menstrual period) |
| Covariate | Hormone replacement therapy | 2814 | Ever used hormone-replacement therapy (HRT) |
| Covariate | Parity | 2734 | Number of live births |
| Covariate | Shift work | 826 | Job involves shift work |
| Covariate | Sleep apnoea | 41280 | ICD-10 diagnosis code G47.3 |
| Exposure | Sleep duration | 1160 | Sleep duration |
| Exposure | Chronotype | 1180 | Morning/evening person (chronotype) |
| Exposure | Nap during day | 1190 | Do you have a nap during the day? |
| Exposure | Daytimde sleepiness | 1220 | How likely are you to doze off or fall asleep during the daytime when you don't mean to? (e.g. when working, reading or driving) |
| Exposure | Insomnia | 1200 | Sleeplessness / insomnia |
| Exposure | Snoring | 1210 | Snoring |

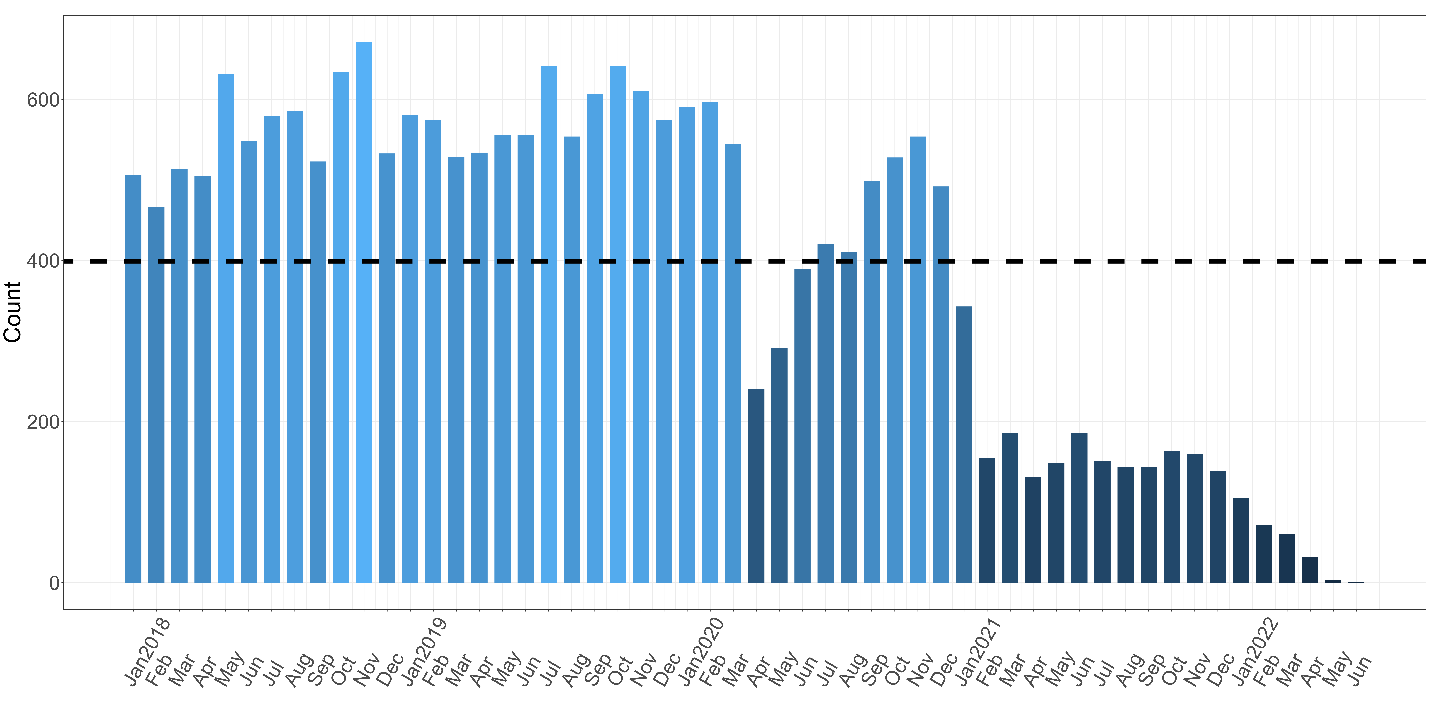

**Figure S2**. Number of cancer cases in UK Biobank recorded from UK registries between 2018 and 2022.

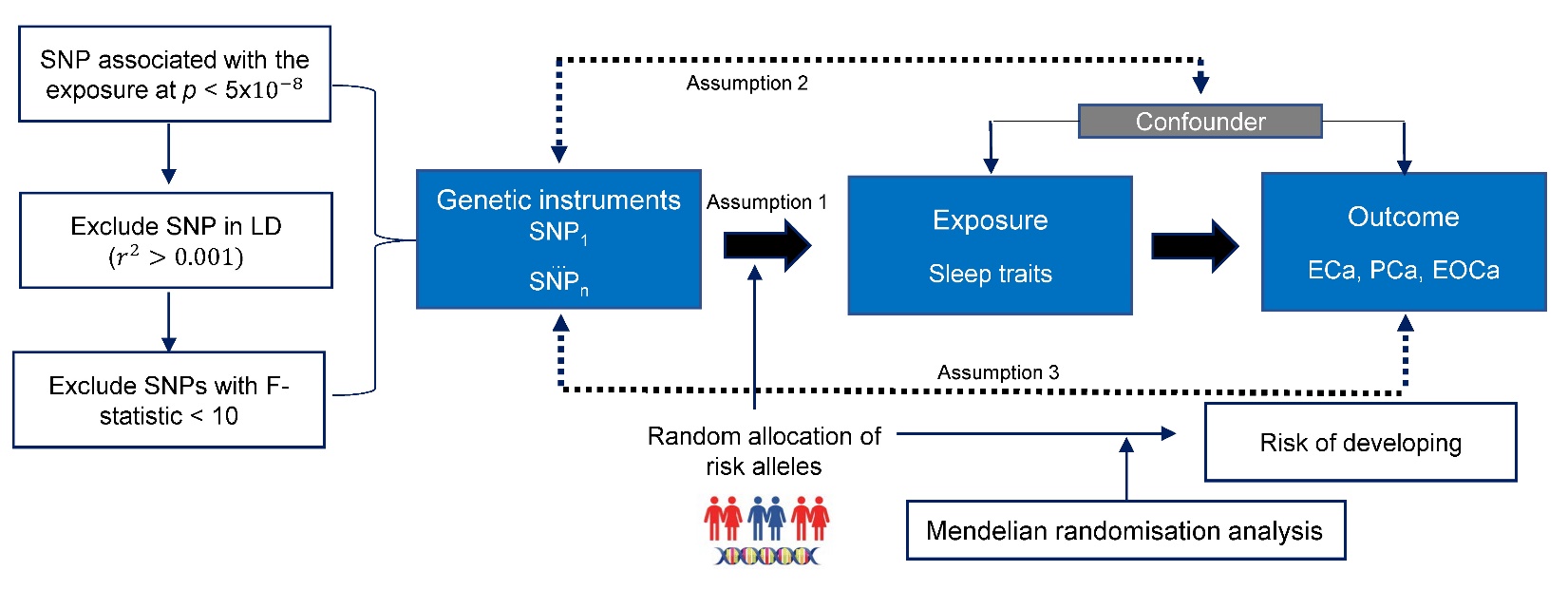

**Figure S3**. Overview and assumptions of the MR design. Dashed lines represent potential pleiotropic or direct causal effects between variables that would violate Mendelian randomisation assumptions. Assumption 1: Genetic variants are associated with the exposure; Assumption 2: Genetic variants are not associated with any confounders; and Assumption 3: Genetic variants influence risk only through the exposure and not through any alternative pathways. The MR design can reduce residual confounding and reverse causality, thereby reinforcing the causal inference of an exposure-outcome association. This is because genetic variants, selected as instrumental variables for studying the effect of modifying the exposure, are randomly allocated at conception and, therefore, less vulnerable to confounding from environmental factors and reverse causation.

**Table S2**. Baseline characteristics of prostate cancer-free stratified by sleep traits in the UK Biobank.

|  | **Snoring** | | **Insomnia** | | | **Sleep duration** | | | **Chronotype** | |
| --- | --- | --- | --- | --- | --- | --- | --- | --- | --- | --- |
|  | **Yes**  **(n=84,735)** | **No**  **(n=92,388)** | **Never/rarely**  **(n=57,358)** | **Sometimes**  **(n=86,003)** | **Usually**  **(n=44,221)** | **<7 h**  **(n=46,492)** | **7-8 h**  **(n=127,317)** | **>8 h**  **(n=13,164)** | **Morning**  **(n=99,189)** | **Evening**  **(n=62, 632)** |
| **Age,** years, mean (SD) | 56.0 (8.0) | 56.0 (8.0) | 56.0 (8.0) | 56.0 (8.0) | 57.0 (8.0) | 55.0 (8.0) | 57.0 (8.0) | 60.0 (7.0) | 57.0 (8.0) | 55.0 (8.0) |
| median (IQR) | 58 (50, 63) | 58 (49,63) | 57 (49, 63) | 58 (50, 63) | 59 (51, 64) | 56 (49, 62) | 58 (50, 63) | 62 (56, 66) | 58 (51, 64) | 56 (48, 62) |
| **BMI,** Kg/m^2^, mean (SD) | 28.6 (4.3) | 27.1 (4.0) | 27.6 (4.1) | 27.8 (4.1) | 28.3 (4.6) | 28.4 (4.5) | 27.6 (4.1) | 28.4 (4.6) | 27.9 (4.2) | 27.9 (4.3) |
| median (IQR) | 28.1 (25.8, 30.9) | 26.6 (24.4, 29.2) | 27.1 (24.9, 29.8) | 27.3 (25.0, 30.0) | 27.7 (25.2, 30.7) | 27.8 (25.3, 30.7)) | 27.1 (24.9, 29.8) | 27.9 (25.3, 30.9) | 27.4 (25.1, 30.1) | 27.3 (25.0, 30.1) |
| Unknown | 349 (0.4) | 525 (0.6) | 270 (0.5) | 400 (0.7) | 292 (0.7) | 276 (0.6) | 562 (0.4) | 114 (0.9) | 461 (0.5) | 356 (0.6) |
| **Smoking status** |  |  |  |  |  |  |  |  |  |  |
| Never | 38,969 (46.0) | 47,188 (51.0) | 30,037 (52.0) | 41,831 (49.0) | 19,421(44.0) | 21,683 (47.0) | 63,899 (50.0) | 5,454 (41.0) | 49,942 (50.0) | 28,010 (45.0) |
| Previous | 34,793 (41.0) | 33,800 (37.0) | 20,427 (36.0) | 33,062 (38.0) | 18,588 (42.0) | 17,621 (38.0) | 48,532 (38.0) | 5,731 (44.0) | 38,682 (39.0) | 24,063 (38.0) |
| Current | 10,718 (13.0) | 11,116 (12.0) | 6,704 (12.0) | 10,824 (13.0) | 6,060 (14.0) | 7,021 (15.0) | 14,491 (11.0) | 1,925 (15.0) | 10,244 (10.0) | 10,398 (17.0) |
| Unknown | 255 (0.3) | 284 (0.3) | 190 (0.3) | 286 (0.3) | 152 (0.3) | 167 (0.4) | 395 (0.3) | 54 (0.4) | 321 (0.3) | 161 (0.3) |
| **Townsend deprivation index,** median (IQR) | -2.4 (-3.8, -0.04) | -2.2 (-3.7, 0.5) | -2.4 (-3.8, 0.02) | -2.2 (-3.7, 0.4) | -2.0 (-3.6, 1.0) | -1.8 (-3.5, 1.1) | -2.4 (-3.8, 0.02) | -1.9 (-3.5, 1.1) | -2.3 (-3.7, 0.1) | -2.0 (-3.6, 0.7) |
| Unknown | 108 (0.1) | 125 (0.1) | 71 (0.1) | 115 (0.1) | 62 (0.1) | 69 (0.1) | 164 (0.1) | 14 (0.1) | 131 (0.1) | 79 (0.1) |
| **Coffee intake**, cup/d |  |  |  |  |  |  |  |  |  |  |
| <1 | 21,178 (25.0) | 25,134 (27.0) | 14,751 (26.0) | 22,288 (26.0) | 12,406 (28.0) | 12,539 (27.0) | 32,765 (26.0) | 3,908 (30.0) | 25,972 (26.0) | 16,608 (27.0) |
| 1-2 | 31,177 (37.0) | 35,217 (38.0) | 21,279 (37.0) | 32,538 (38.0) | 16,307 (37.0) | 15,937 (34.0) | 49,096 (39.0) | 4,960 (38.0) | 38,277 (39.0) | 22,330 (36.0) |
| 3-4 | 19,928 (24.0) | 20,370 (22.0) | 13,240 (23.0) | 19,779 (23.0) | 9,414 (21.0) | 10,246 (22.0) | 29,424 (23.0) | 2,662 (20.0) | 22,294 (22.0) | 14,229 (23.0) |
| 5-6 | 9,042 (11.0) | 8,485 (9.2) | 5,920 (10.0) | 8,278 (9.6) | 4,264 (9.6) | 5,339 (11.0) | 11,993 (9.4) | 1,080 (8.2) | 9,229 (9.3) | 6,793 (11.0) |
| >6 | 3,292 (3.9) | 3,1027 (3.3) | 2,089 (3.6) | 2,947 (3.4) | 1,729 (3.9) | 2,331 (5.0) | 3,878 (3.0) | 520 (4.0) | 3,285 (3.3) | 2,583 (4.1) |
| Unknown | 118 (0.1) | 155 (0.2) | 79 (0.1) | 173 (0.2) | 101 (0.2) | 100 (0.2) | 161 (0.1) | 34 (0.3) | 132 (0.1) | 89 (0.1) |
| **Tea intake**, cup/d |  |  |  |  |  |  |  |  |  |  |
| <1 | 15,419 (18.0) | 15,784 (17.0) | 10,651 (19.0) | 14,728 (17.0) | 7,855 (18.07) | 8,887 (19.0) | 21,856 (17.0) | 2,359 (18.0) | 16,705 (17.0) | 11,944 (19.0) |
| 1-2 | 19,240 (23.0) | 21,207 (23.0) | 13,437 (23.0) | 19,653 (23.0) | 9,631 (22.0) | 10,199 (22.0) | 29,665 (23.0) | 2,774 (21.0) | 22,630 (23.0) | 14,216 (23.0) |
| 3-4 | 23,638 (28.0) | 26,563 (29.0) | 15,815 (28.0) | 24,900 (29.0) | 12,251 (28.0) | 12,323 (27.0) | 36,891 (29.0) | 3,632 (28.0) | 28,761 (29.0) | 16,875 (27.0) |
| 5-6 | 17,304 (20.0) | 18,867 (20.5) | 11,429 (20.0) | 17,713 (21.0) | 9,067 (21.0) | 9,364 (20.0) | 25,962 (20.0) | 2,755 (21.0) | 20,597 (21.0) | 12,411 (20.0) |
| >6 | 9,003 (11.0) | 9,802 (11.0) | 5,942 (10.0) | 8,833 (10.0) | 5,299 (12.0) | 5,601 (12.0) | 12,789 (10.0) | 1,612 (12.0) | 10,369 (10.0) | 7,086 (11.0) |
| Unknown | 131 (0.2) | 165 (0.2) | 84 (0.1) | 176 (0.2) | 118 (0.3) | 118 (0.2) | 154 (0.1) | 32 (0.2) | 127 (0.1) | 100 (0.2) |
| **Total MET-h/w,** median (IQR) | 29 (13, 60) | 33 (15, 66) | 32 (15, 65) | 31 (14, 63) | 29 (12, 61) | 31 (13, 68) | 31 (14, 62) | 28 (11, 60) | 33 (15, 67) | 27 (12, 56) |
| Unknown | 13,144 (15.5) | 13,808 (14.9) | 8,015 (14.0) | 14,030 (16.3) | 7,465 (16.9) | 7,962 (17.1) | 18,776 (14.7) | 2,249 (17.1) | 13,345 (13.5) | 9,096 (14.5) |
| **Prostate-specific antigen test history** | 23,956 (28.0) | 24,747 (27.0) | 13,971 (24.0) | 22,751 (26.0) | 14,278 (32.0) | 11,800 (25.0) | 34,918 (27.0) | 4,195 (32.0) | 28,710 (29.0) | 15,648 (25.0) |
| Unknown | 4,508 (5.3) | 4,594 (5.0) | 2,475 (4.3) | 4,592 (5.3) | 2,868 (6.5) | 2,784 (6.0) | 6,250 (4.9) | 798 (6.1) | 4,888 (4.9) | 3,354 (5.4) |
| **Education** |  |  |  |  |  |  |  |  |  |  |
| College or University | 24,572 (29.0) | 29,517 (31.9) | 19,135 (33.4) | 25,795 (30.0) | 11,872 (27.7) | 12,313 (26.5) | 41,604 (32.7) | 2,870 (21.8) | 29,694 (29.9) | 19,778 (31.6) |
| A levels/AS levels | 8,224 (9.7) | 8,558 (9.3) | 5,745 (10.0) | 8,025 (9.3) | 3,959 (7.2) | 4,295 (9.2) | 12,412 (9.7) | 1,006 (3.7) | 8,872 (8.9) | 6,536 (10.4) |
| O levels/GCSEs | 15,368 (18.1) | 15,163 (16.4) | 9,552 (16.7) | 14,981 (17.4) | 7,646 (13.8) | 8,378 (18.0) | 21,683 (17.0) | 2,048 (12.1) | 16,722 (16.9) | 11,290 (18.0) |
| CSEs | 4,600 (5.4) | 4,524 (4.9) | 2,899 (5.1) | 4,628 (5.4) | 2,265 (4.6) | 2,961 (6.4) | 6,217 (4.9) | 571 (6.5) | 5,046 (5.1) | 3,406 (5.4) |
| NVQ, HND, HNC | 7,354 (8.7) | 7,184 (7.8) | 4,497 (7.8) | 7,182 (8.4) | 3,698 (5.6) | 3,786 (8.1) | 10,291 (8.1) | 1,262 (6.1) | 8,608 (8.7) | 4,915 (7.8) |
| Other | 15,869 (18.7) | 17,797 (19.3) | 9,486 (16.5) | 16,416 (19.1) | 10,206 (26.2) | 9,862 (21.2) | 21,936 (17.2) | 3,986 (46.6) | 20,607 (20.8) | 10,731 (17.1) |
| Unknown | 8,748 (10.3) | 9,645 (10.4) | 6,044 (10.5) | 8,976 (10.4) | 4,575 (14.9) | 4,897 (10.5) | 13,174 (10.3) | 1,421 (16.4) | 9,640 (9.7) | 5,976 (9.5) |

**Table S3**. Baseline characteristics of endometrial cancer-free stratified by sleep traits in the UK Biobank.

|  | **Snoring** | | **Insomnia** | | | **Sleep duration** | | | **Chronotype** | |
| --- | --- | --- | --- | --- | --- | --- | --- | --- | --- | --- |
|  | **Yes**  **(n=46,481)** | **No**  **(n=121,676)** | **Never/rarely**  **(n=36,611)** | **Sometimes**  **(n=90,527)** | **Usually**  **(n=55,375)** | **<7 h**  **(n=40,677)** | **7-8 h**  **(n=127,004)** | **>8 h**  **(n=13,818)** | **Morning**  **(n=104,079)** | **Evening**  **(n=61,846)** |
| **Age,** years, mean (SD) | 56.0 (8.0) | 55.0 (8.0) | 53.0 (8.0) | 56.0 (8.0) | 57.0 (8.0) | 56.0 (8.0) | 55.0 (8.0) | 56.0 (8.0) | 56.0 (8.0) | 55.0 (8.0) |
| median (IQR) | 57 (50, 62) | 56 (48, 62) | 52 (46, 61) | 57 (49, 62) | 57 (51, 62) | 57 (51, 62) | 56 (48, 62) | 58 (49, 63) | 57 (50, 62) | 55 (48, 62) |
| **BMI,** Kg/m^2^, mean (SD) | 28.5 (5.5) | 26.2 (4.7) | 26.3 (4.9) | 26.6 (4.9) | 27.3 (5.4) | 27.3 (5.5) | 26.5 (4.9) | 27.6 (5.4) | 26.6 (5.0) | 27.1 (5.2) |
| median (IQR) | 27.5 (24.5, 31.4) | 25.2 (22.8, 28.3) | 25.4 (22.9, 28.8) | 25.7 (23.2, 29.1) | 26.3 (23.5, 30.0) | 26.3 (23.5, 30.1) | 25.6 (23.1, 28.9) | 26.7 (23.8, 30.4) | 25.7 (23.1, 29.1) | 26.1 (23.4, 29.7) |
| Unknown | 160 (0.3) | 466 (0.4) | 139 (0.4) | 315 (0.3) | 252 (0.5) | 191 (0.5) | 416 (0.3) | 92 (0.7) | 359 (0.3) | 260 (0.4) |
| **Smoking status** |  |  |  |  |  |  |  |  |  |  |
| Never | 25,803 (55.5) | 74,670 (61.3) | 23,090 (63.1) | 54,698 (60.4) | 30,847 (55.7) | 22,931 (56.4) | 77,201 (60.8) | 7,912 (57.3) | 64,514 (62.0) | 33,755 (54.6) |
| Previous | 15,653 (33.7) | 37,295 (30.7) | 10,324 (28.2) | 28,083 (31.0) | 18,959 (34.2) | 13,259 (32.6) | 39,398 (31.0) | 4,429 (32.1) | 32,169 (30.9) | 20,347 (32.9) |
| Current | 4,873 (10.5) | 9,389 (7.7) | 3,094 (8.5) | 7,496 (8.3) | 5,368 (9.7) | 4,317 (10.6) | 10,087 (7.9) | 1,434 (10.4) | 7,085 (6.8) | 7,591 (12.3) |
| Unknown | 152 (0.3) | 322 (0.3) | 103 (0.3) | 250 (0.3) | 201 (0.4) | 170 (0.4) | 318 (0.3) | 43 (0.3) | 311 (0.3) | 153 (0.2) |
| **Townsend deprivation index,** median (IQR) | -2.2 (-3.7, 0.2) | -2.4 (-3.7, 0.02) | -2.3 (-3.7, 0.09) | -2.3 (-3.7, 0.1) | -2.2 (-3.6, 0.4) | -2.0 (-3.6, 0.7) | -2.4 (-3.7, -0.0) | -2.2 (-3.6, 0.4) | -2.3 (-3.7, 0.05) | -2.1 (-3.6, 0.4) |
| Unknown | 50 (0.1) | 138 (0.1) | 36 (<0.1) | 106 (0.1) | 65 (0.1) | 54 (0.1) | 133 (0.1) | 19 (0.1) | 108 (0.1) | 81 (0.1) |
| **Coffee intake**, cup/d |  |  |  |  |  |  |  |  |  |  |
| <1 | 13,606 (29.2) | 36,726 (30.2) | 11,126 (30.4) | 26,415 (29.2) | 17,027 (30.7) | 12,515 (30.8) | 37,230 (29.3) | 4,453 (32.2) | 30,781 (29.6) | 18,697 (30.2) |
| 1-2 | 18,001 (38.7) | 49,243 (40.5) | 14,559 (39.8) | 36,879 (40.7) | 21,386 (38.6) | 15,253 (37.5) | 51,934 (40.9) | 5,310 (38.4) | 42,873 (41.2) | 23,519 (38.0) |
| 3-4 | 9,665 (20.8) | 24,290 (20.0) | 7,156 (19.5) | 18,585 (20.5) | 11,053 (20.0) | 8,174 (20.1) | 25,791 (20.3) | 2,663 (19.3) | 20,765 (20.0) | 12,649 (20.5) |
| 5-6 | 3,853 (8.3) | 8,637 (7.1) | 2,838 (7.8) | 6,530 (7.2) | 4,294 (7.8) | 3,381 (8.3) | 9,183 (7.2) | 1,025 (7.4) | 7,289 (7.0) | 5,135 (8.3) |
| >6 | 1,295 (2.8) | 2,667 (2.2) | 890 (2.4) | 2,005 (2.2) | 1,527 (2.8) | 1,283 (3.2) | 2,757 (2.2) | 346 (2.5) | 2,267 (2.2) | 1,770 (2.9) |
| Unknown | 61 (0.1) | 113 (0.1) | 42 (0.1) | 113 (0.1) | 88 (0.2) | 71 (0.2) | 109 (0.1) | 21 (0.2) | 104 (0.1) | 76 (0.1) |
| **Tea intake**, cup/d |  |  |  |  |  |  |  |  |  |  |
| <1 | 8,779 (18.9) | 21,817 (17.9) | 6,831 (18.7) | 15,901 (17.6) | 10,742 (19.4) | 8,258 (20.3) | 22,421 (17.7) | 2,582 (18.7) | 18,439 (17.7) | 11,991 (19.4) |
| 1-2 | 9,662 (20.8) | 26,979 (22.2) | 8,309 (22.7) | 19,835 (21.9) | 11,557 (20.9) | 8,405 (20.7) | 28,243 (22.2) | 2,865 (20.7) | 22,760 (21.9) | 13,422 (21.7) |
| 3-4 | 13,601 (29.3) | 36,706 (30.2) | 10,614 (29.0) | 27,719 (30.6) | 15,876 (28.7) | 11,286 (27.7) | 38,733 (30.5) | 3,939 (28.5) | 31,823 (30.6) | 17,393 (28.1) |
| 5-6 | 9,739 (21.1) | 24,760 (20.3) | 7,267 (19.8) | 18,714 (20.7) | 11,451 (20.7) | 8,355 (20.5) | 26,004 (20.5) | 2,891 (20.9) | 21,456 (20.6) | 12,598 (20.4) |
| >6 | 4,619 (9.9) | 11,235 (9.2) | 3,549 (9.7) | 8,209 (9.1) | 5,621 (10.2) | 4,293 (10.6) | 11,456 (9.0) | 1,510 (10.9) | 9,457 (9.1) | 6,344 (10.3) |
| Unknown | 81 (0.2) | 179 (0.2) | 41 (0.1) | 149 (0.2) | 128 (0.2) | 80 (0.2) | 147 (0.1) | 31 (0.2) | 144 (0.1) | 98 (0.2) |
| **Menopause** | 32,278 (69.4) | 76,714 (63.0) | 18,720 (51.1) | 59,949 (66.2) | 40,707 (73.5) | 29,046 (71.4) | 80,236 (63.2) | 9,339 (67.6) | 70,590 (67.8) | 38,188 (61.7) |
| Unknown | 2,576 (5.5) | 6,061 (5.0) | 1,850 (5.1) | 4,502 (5.0) | 3,077 (5.6) | 2,238 (5.5) | 6,450 (5.1) | 663 (4.8) | 5,030 (4.8) | 3,440 (5.6) |
| **Hormone replacement therapy** | 15,910 (34.2) | 36,267 (29.8) | 8,055 (22.0) | 28,027 (31.0) | 20,991 (37.9) | 14,214 (35.0) | 37,551 (29.7) | 4,902 (35.5) | 33,637 (32.3) | 18,649 (30.2) |
| Unknown | 145 (0.3) | 266 (0.2) | 83 (0.2) | 238 (0.3) | 170 (0.3) | 122 (0.3) | 312 (0.2) | 43 (0.3) | 39 (0.2) | 168 (0.3) |
| **Total MET-h/w,** median (IQR) | 26 (12, 53) | 30 (15, 58) | 30 (15, 58) | 29 (14, 56) | 28 (12, 56) | 29 (13, 59) | 29 (14, 56) | 27 (12, 56) | 31 (15, 60) | 26 (12, 51) |
| Unknown | 10,613 (22.8) | 24,738 (20.3) | 6,403 (17.5) | 19,746 (21.8) | 13,223 (23.9) | 9,600 (23.6) | 25,790 (20.3) | 3,209 (23.2) | 20,349 (19.6) | 12,984 (21.0) |
| **Parity** |  |  |  |  |  |  |  |  |  |  |
| 0 | 7,918 (17.0) | 24,135 (19.8) | 8,249 (22.5) | 17,862 (19.7) | 10,535 (19.0) | 8,146 (20.2) | 25,662 (20.2) | 2,672 (19.3) | 19,335 (18.6) | 14,029 (22.7) |
| ≥1 | 38,532 (82.9) | 97,479 (80.1) | 28,338 (77.4) | 72,609 (80.2) | 44,794 (80.9) | 32,499 (79.9) | 101,266 (79.7) | 11,137 (80.6) | 84,677 (81.4) | 47,778 (77.3) |
| Unknown | 31 (0.1) | 62 (0.1) | 24 (0.1) | 56 (0.1) | 46 (0.1) | 32 (0.1) | 76 (0.1) | 9 (0.1) | 67 (0.1) | 39 (0.1) |
| **Education** |  |  |  |  |  |  |  |  |  |  |
| College or University | 12,300 (26.5) | 39,112 (32.1) | 13,320 (36.4) | 27,755 (30.7) | 14,327 (25.9) | 10,662 (26.2) | 41,352 (32.6) | 3,285 (23.8) | 30,859 (29.6) | 19,794 (32.0) |
| A levels/AS levels | 5,035 (10.8) | 14,077 (11.6) | 4,584 (12.5) | 10,238 (11.3) | 5,868 (10.6) | 4,249 (10.4) | 14,986 (11.8) | 1,397 (10.1) | 11,514 (11.1) | 7,463 (12.1) |
| O levels/GCSEs | 10,368 (22.3) | 25,253 (20.8) | 7,413 (20.2) | 19,036 (21.0) | 12,100 (21.9) | 8,688 (21.4) | 26,755 (21.1) | 2,916 (21.1) | 22,202 (21.3) | 13,216 (21.4) |
| CSEs | 2,591 (5.6) | 5,730 (4.7) | 1,695 (4.6) | 4,361 (4.8) | 2,933 (5.3) | 2,137 (5.3) | 5,993 (4.7) | 802 (5.8) | 5,163 (5.0) | 3,083 (5.0) |
| NVQ, HND, HNC | 2,007 (4.3) | 4,203 (3.5) | 1,171 (3.2) | 3,291 (3.6) | 2,354 (4.3) | 1,822 (4.5) | 4,319 (3.4) | 802 (4.5) | 4,122 (4.0) | 2,198 (3.6) |
| Other | 9,449 (20.3) | 20,904 (17.2) | 4,768 (13.0) | 16,536 (18.2) | 12,048 (21.8) | 8,869 (21.8) | 20,677 (16.3) | 3,384 (24.5) | 20,255 (19.5) | 10,223 (16.5) |
| Unknown | 4,731 (10.2) | 12,397 (10.2) | 3,660 (10.0) | 9,310 (10.3) | 5,745 (10.4) | 4,250 (10.4) | 12,922 (10.2) | 1,415 (10.2) | 9,964 (9.6) | 5,869 (9.5) |

**Table S4.** Baseline characteristics of epithelial ovarian cancer-free stratified by sleep traits in the UK Biobank.

|  | **Snoring** | | **Insomnia** | | | **Sleep duration** | | | **Chronotype** | |
| --- | --- | --- | --- | --- | --- | --- | --- | --- | --- | --- |
|  | **Yes**  **(n=46,674)** | **No**  **(n=121,861)** | **Never/rarely**  **(n=36,660)** | **Sometimes**  **(n=90,730)** | **Usually**  **(n=55,523)** | **<7 h**  **(n=40,770)** | **7-8 h**  **(n=127,267)** | **>8 h**  **(n=13,855)** | **Morning**  **(n=104,304)** | **Evening**  **(n=62,002)** |
| **Age,** years, mean (SD) | 56.0 (8.0) | 55.0 (8.0) | 53.0 (8.0) | 56.0 (8.0) | 57.0 (7.5) | 56.0 (8.0) | 55.0 (8.0) | 56.0 (8.0) | 56.0 (8.0) | 55.0 (8.0) |
| median (IQR) | 57 (50, 62) | 56 (48, 62) | 52 (46, 61) | 57 (49, 62) | 57 (51, 62) | 57 (51, 62) | 56 (48, 62) | 58 (49, 63) | 57 (50, 62) | 55 (48, 62) |
| **BMI,** Kg/m^2^, mean (SD) | 28.5 (5.5) | 26.1 (4.7) | 26.4 (4.9) | 26.6 (4.9) | 27.3 (5.4) | 27.4 (5.5) | 26.5 (4.9) | 27.6 (5.4) | 26.6 (5.0) | 27.1 (5.2) |
| median (IQR) | 27.5 (24.6, 31.4) | 25.2 (22.8, 28.4) | 25.4 (22.9, 28.8) | 25.7 (23.2, 29.1) | 26.3 (23.5, 30.0) | 26.3 (23.5, 30.1) | 25.6 (23.1, 28.9) | 26.7 (23.8, 30.4) | 25.7 (23.1, 29.1) | 26.1 (23.4, 29.7) |
| Unknown | 162 (0.3) | 466 (0.4) | 139 (0.4) | 316 (0.3) | 254 (0.5) | 191 (0.5) | 420 (0.3) | 91 (0.7) | 361 (0.3) | 261 (0.4) |
| **Smoking status** |  |  |  |  |  |  |  |  |  |  |
| Never | 25,945 (55.6) | 74,826 (61.4) | 23,129 (63.1) | 54,867 (60.5) | 30,962 (55.8) | 23,016 (56.4) | 77,406 (60.8) | 7,940 (57.3) | 64,703 (62.0) | 33,878 (54.6) |
| Previous | 15,706 (33.7) | 37,324 (30.6) | 10,340 (28.2) | 28,121 (31.0) | 18,984 (34.2) | 13,268 (32.5) | 39,461 (31.0) | 4,435 (32.0) | 32,197 (30.9) | 20,386 (32.9) |
| Current | 4,870 (10.4) | 9,387 (7.7) | 3,087 (8.4) | 7,490 (8.3) | 5,376 (9.7) | 4,317 (10.6) | 10,079 (7.9) | 1,436 (10.4) | 7,090 (6.8) | 7,585 (12.2) |
| Unknown | 153 (0.3) | 324 (0.3) | 104 (0.3) | 252 (0.3) | 201 (0.4) | 169 (0.4) | 321 (0.3) | 44 (0.3) | 314 (0.3) | 153 (0.2) |
| **Townsend deprivation index,** median (IQR) | -2.2 (-3.7, 0.2) | -2.3 (-3.7, 0.02) | -2.3 (-3.7, 0.1) | -2.3 (-3.7, 0.1) | -2.2 (-3.6, 0.4) | -2.0 (-3.6, 0.7) | -2.4 (-3.7, 0.0) | -2.2 (-3.6, 0.5) | -2.3 (-3.7, 0.05) | -2.1 (-3.6, 0.4) |
| Unknown | 50 (0.1) | 138 (0.1) | 36 (<0.1) | 106 (0.1) | 77 (0.1) | 54 (0.1) | 133 (0.1) | 19 (0.1) | 108 (0.1) | 81 (0.1) |
| **Coffee intake**, cup/d |  |  |  |  |  |  |  |  |  |  |
| <1 | 13,673 (29.3) | 36,774 (30.2) | 11,138 (30.4) | 26,479 (29.2) | 17,075 (30.7) | 12,547 (30.8) | 37,300 (29.3) | 4,470 (32.3) | 30,854 (29.6) | 18,739 (30.2) |
| 1-2 | 18,072 (38.7) | 49,323 (40.5) | 14,578 (39.8) | 36,975 (40.7) | 21,422 (38.9) | 15,284 (37.4) | 52,033 (40.9) | 5,326 (38.4) | 42,955 (41.2) | 23,583 (38.0) |
| 3-4 | 9,701 (20.8) | 24,349 (20.0) | 7,173 (19.6) | 18,623 (20.5) | 11,100 (20.0) | 8,200 (20.1) | 25,865 (20.3) | 2,667 (19.2) | 20,817 (20.0) | 12,694 (20.5) |
| 5-6 | 3,867 (8.3) | 8,639 (7.1) | 2,837 (7.7) | 6,536 (7.2) | 4,309 (7.8) | 3,386 (8.3) | 9,196 (7.2) | 1,027 (7.4) | 7,305 (7.0) | 5,140 (8.3) |
| >6 | 1,298 (2.8) | 2,663 (2.2) | 891 (2.4) | 2,003 (2.2) | 1,530 (2.8) | 1,281 (3.1) | 2,763 (2.2) | 344 (2.5) | 2,266 (2.2) | 1,772 (2.9) |
| Unknown | 63 (0.1) | 113 (0.1) | 43 (0.1) | 114 (0.1) | 87 (0.2) | 72 (0.2) | 110 (0.1) | 21 (0.2) | 107 (0.1) | 74 (0.1) |
| **Tea intake**, cup/d |  |  |  |  |  |  |  |  |  |  |
| <1 | 8,817 (18.9) | 21,838 (17.9) | 6,841 (18.7) | 15,938 (17.6) | 10,771 (19.4) | 8,265 (20.3) | 22,481 (17.7) | 2,591 (18.7) | 18,499 (17.7) | 12,003 (19.4) |
| 1-2 | 9,708 (20.8) | 27,008 (22.2) | 8,336 (22.7) | 19,869 (21.9) | 11,578 (20.9) | 8,416 (20.6) | 28,312 (22.5) | 2,864 (20.7) | 22,789 (21.8) | 13,464 (21.7) |
| 3-4 | 13,653 (29.3) | 36,778 (30.2) | 10,624 (29.0) | 27,790 (30.6) | 15,916 (28.7) | 11,326 (27.8) | 38,794 (30.5) | 3,958 (28.6) | 31,879 (30.6) | 17,449 (28.1) |
| 5-6 | 9,775 (20.9) | 24,807 (20.4) | 7,267 (19.8) | 18,759 (20.7) | 11,496 (20.7) | 8,386 (20.6) | 26,060 (20.5) | 2,892 (20.9) | 21,514 (20.6) | 12,627 (20.4) |
| >6 | 4,639 (9.9) | 11,250 (9.2) | 3,550 (9.7) | 8,225 (9.1) | 5,634 (10.1) | 4,297 (10.5) | 11,471 (9.0) | 1,519 (11.0) | 9,478 (9.1) | 6,361 (10.3) |
| Unknown | 82 (0.2) | 180 (0.1) | 42 (0.1) | 149 (0.2) | 128 (0.2) | 80 (0.2) | 149 (0.1) | 31 (0.2) | 145 (0.1) | 98 (0.2) |
| **Menopause** | 32,432 (69.5) | 76,880 (63.1) | 18,753 (51.2) | 60,126 (66.3) | 40,831 (73.5) | 29,120 (71.4) | 80,460 (63.2) | 9,372 (67.6) | 70,785 (67.9) | 38,308 (61.8) |
| Unknown | 2,585 (5.5) | 6,068 (4.5) | 1,856 (5.1) | 4,506 (5.0) | 3,086 (5.6) | 2,244 (5.5) | 6,458 (5.1) | 665 (4.8) | 5,042 (4.8) | 3,448 (5.6) |
| **Hormone replacement therapy** | 15,963 (34.2) | 36,334 (29.8) | 8,068 (22.0) | 28,089 (31.0) | 21,041 (37.9) | 14,236 (34.9) | 37,643 (29.6) | 4,910 (35.4) | 33,715 (32.3) | 18,690 (30.1) |
| Unknown | 145 (0.3) | 267 (0.2) | 83 (0.2) | 240 (0.3) | 168 (0.3) | 122 (0.3) | 312 (0.2) | 43 (0.3) | 240 (0.2) | 166 (0.3) |
| **Total MET-h/w,** median (IQR) | 26 (12, 53) | 30 (15, 58) | 30 (15, 58) | 29 (14, 56) | 28 (12, 56) | 29 (13, 59) | 29 (14, 56) | 27 (12, 56) | 31 (15, 60) | 26 (12, 51) |
| Unknown | 10,661 (22.8) | 24,774 (20.3) | 6,399 (17.5) | 19,808 (21.8) | 13,254 (23.9) | 9,631 (23.6) | 25,832 (20.3) | 3,222 (23.3) | 20,389 (19.5) | 13,031 (21.0) |
| **Parity** |  |  |  |  |  |  |  |  |  |  |
| 0 | 7,958 (17.1) | 24,182 (19.8) | 8,263 (22.5) | 17,921 (19.8) | 10,570 (19.0) | 8,171 (20.0) | 25,734 (20.2) | 2,684 (19.4) | 19,380 (18.9) | 14,078 (22.7) |
| ≥1 | 38,684 (82.9) | 97,617 (80.1) | 28,373 (77.4) | 72,753 (80.2) | 44,906 (80.9) | 37,878 (79.9) | 101,457 (79.7) | 11,162 (80.6) | 84,856 (81.4) | 47,885 (77.2) |
| Unknown | 32 (0.1) | 62 (0.1) | 24 (0.1) | 56 (0.1) | 47 (0.1) | 33 (0.1) | 76 (0.1) | 9 (0.1) | 68 (0.1) | 39 (0.1) |
| **Education** |  |  |  |  |  |  |  |  |  |  |
| College or University | 12,351 (26.4) | 39,172 (32.1) | 13,345 (36.4) | 27,816 (30.7) | 14,363 (25.9) | 10,693 (26.2) | 41,433 (32.6) | 3,293 (23.8) | 30,920 (29.6) | 19,846 (32.0) |
| A levels/AS levels | 5,060 (10.8) | 14,079 (11.6) | 4,579 (12.5) | 10,252 (11.3) | 5,887 (10.6) | 4,257 (10.4) | 15,006 (11.8) | 1,399 (10.1) | 11,525 (11.0) | 7,473 (12.1) |
| O levels/GCSEs | 10,404 (22.3) | 25,270 (20.7) | 7,415 (20.2) | 19,065 (21.0) | 12,120 (21.8) | 8,705 (21.4) | 26,788 (21.0) | 2,916 (21.0) | 22,232 (21.3) | 13,244 (21.4) |
| CSEs | 2,599 (5.6) | 5,743 (4.7) | 1,696 (4.6) | 4,376 (4.8) | 2,939 (5.3) | 2,134 (5.2) | 6,012 (4.7) | 807 (5.8) | 5,173 (5.0) | 3,095 (5.0) |
| NVQ, HND, HNC | 2,016 (4.3) | 4,210 (3.5) | 1,176 (3.2) | 3,296 (3.6) | 2,364 (4.3) | 1,829 (4.3) | 6,012 (3.4) | 621 (4.5) | 4,138 (4.0) | 2,205 (3.6) |
| Other | 9,487 (20.3) | 20,956 (17.2) | 4,780 (13.0) | 16,585 (18.3) | 12,078 (21.8) | 8,883 (21.8) | 20,739 (16.3) | 3,397 (24.5) | 20,317 (19.5) | 10,247 (16.5) |
| Unknown | 4,757 (10.2) | 12,431 (10.2) | 3,669 (10.0) | 9,340 (10.3) | 5,772 (10.4) | 4,269 (10.5) | 12,960 (10.2) | 1,422 (10.3) | 9,999 (9.6) | 5,892 (9.5) |

**Table S5**. Associations of sleep traits with reproductive system cancer incidence in the UK Biobank population

|  | **Events** | **Adjusted HR^a^**  **(95% CI)** | **P-value** | **Adjusted HR^b,c^**  **(95% CI)** | **P-value** |
| --- | --- | --- | --- | --- | --- |
| **Prostate cancer** | | | | | |
| **Sleep duration** | |  |  |  |  |
| <7 h | 1,921 | 0.98 (0.93, 1.03) | 0.464 | 0.99 (0.93, 1.05) | 0.631 |
| 7-8 h | 5,968 | Reference |  | Reference |  |
| >8 h | 685 | 0.92 (0.85, 1.00) | 0.045 | 0.90 (0.82, 0.99) | 0.037 |
| Per 1h |  | 1.00 (0.98, 1.02) | 0.774 | 0.99 (0.96, 1.01) | 0.295 |
| **Chronotype** | |  |  |  |  |
| Morning | 4,776 | Reference |  | Reference |  |
| Evening | 2,635 | 1.00 (0.95, 1.04) | 0.834 | 1.00 (0.95, 1.06) | 0.871 |
| **Chronotype** |  |  |  |  |  |
| Definitely morning | 2,038 | Reference |  | Reference |  |
| More morning | 2,738 | 1.00 (0.94, 1.06) | 0.981 | 1.00 (0.93, 1.06) | 0.885 |
| More evening | 2,031 | 1.00 (0.94, 1.06) | 0.864 | 1.00 (0.93, 1.07) | 0.991 |
| Definitely evening | 604 | 1.00 (0.91, 1.09) | 0.939 | 1.01 (0.91, 1.02) | 0.845 |
| Mixed | 1,104 | 0.99 (0.92, 1.06) | 0.730 | 1.02 (0.94, 1.12) | 0.619 |
| **Insomnia** | |  |  |  |  |
| Never/rarely | 2,442 | Reference |  | Reference |  |
| Sometimes | 3,992 | 1.05 (1.00, 1.11) | 0.043 | 1.05 (0.99, 1.12) | 0.089 |
| Usually | 2,167 | 1.06 (1.00, 1.12) | 0.052 | 1.03 (0.96, 1.10) | 0.414 |
| No (ref) vs Yes | 6,159 | 1.06 (1.01, 1.11) | 0.024 | 1.04 (0.99-1.10) | 0.121 |
| **Daytime sleepiness** |  |  |  |  |  |
| Never/rarely | 6,210 | Reference |  | Reference |  |
| Sometimes | 2,099 | 1.00 (0.89, 1.14) | 0.939 | 0.98 (0.93, 1.04) | 0.562 |
| Often | 268 | 1.00 (0.95, 1.05) | 0.947 | 0.96 (0.82, 1.11) | 0.591 |
| Per category increase | 8,577 | 1.00 (0.96, 1.04) | 0.995 | 0.98 (0.94, 1.03) | 0.459 |
| **Daytime napping** |  |  |  |  |  |
| Never/rarely | 4,060 | Reference |  | Reference |  |
| Sometimes | 3,821 | 0.97 (0.93, 1.02) | 0.212 | 0.97 (0.92, 1.02) | 0.240 |
| Usually | 722 | 0.92 (0.85, 1.00) | 0.058 | 0.99 (0.90, 1.08) | 0.801 |
| Per category increase | 8,602 | 0.97 (0.93, 1.00) | 0.046 | 0.98 (0.95,1.02) | 0.415 |
| **Snoring** | |  |  |  |  |
| No | 4,182 | Reference |  | Reference |  |
| Yes | 3,986 | 1.05 (1.01, 1.10) | 0.021 | 1.04 (0.98, 1.09) | 0.178 |
| * |  | 1.05 (1.01, 1.10) | 0.019 | 1.04 (0.98, 1.09) | 0.166 |
| ** |  | 1.05 (1.01, 1.10) | 0.019 | 1.04 (0.98, 1.09) | 0.177 |
| *** |  | 1.05 (1.01, 1.10) | 0.018 | 1.04 (0.99, 1.09) | 0.160 |
| **Endometrial cancer** | | | | | |
| **Sleep duration** | |  |  |  |  |
| <7 h | 258 | 0.96 (0.83, 1.11) | 0.556 | 0.89 (0.74, 1.07) | 0.204 |
| 7-8 h | 710 | Reference |  | Reference |  |
| >8 h | 99 | 1.03 (0.84, 1.28) | 0.757 | 0.93 (0.71, 1.23) | 0.618 |
| Per 1h |  | 1.05 (1.00, 1.10) | 0.077 | 1.04 (0.98, 1.11) | 0.223 |
| **Chronotype** | |  |  |  |  |
| Morning | 605 | Reference |  | Reference |  |
| Evening | 384 | 1.06 (0.93, 1.20) | 0.393 | 1.07 (0.91, 1.25) | 0.441 |
| **Chronotype** |  |  |  |  |  |
| Definitely morning | 265 | Reference |  | Reference |  |
| More morning | 340 | 1.03 (0.88, 1.21) | 0.727 | 0.99 (0.81, 1.20) | 0.884 |
| More evening | 290 | 1.09 (0.92, 1.28) | 0.332 | 1.09 (0.88, 1.33) | 0.435 |
| Definitely evening | 94 | 1.04 (0.82, 1.32) | 0.756 | 0.96 (0.71, 1.30) | 0.802 |
| Mixed | 79 | 0.95 (0.74, 1.23) | 0.707 | 0.90 (0.64, 1.25) | 0.519 |
| **Insomnia** | |  |  |  |  |
| Never/rarely | 193 | Reference |  | Reference |  |
| Sometimes | 544 | 0.99 (0.84, 1.16) | 0.870 | 1.07 (0.87, 1.31) | 0.523 |
| Usually | 342 | 0.88 (0.74, 1.06) | 0.174 | 1.02 (0.82, 1.28) | 0.857 |
| No (ref) vs Yes | 886 | 0.94 (0.81, 1.10) | 0.476 | 1.00 (0.90, 1.11) | 0.988 |
| **Daytime sleepiness** |  |  |  |  |  |
| Never/rarely | 822 | Reference |  | Reference |  |
| Sometimes | 219 | 0.88 (0.76, 1.02) | 0.100 | 0.78 (0.64, 0.96) | 0.016 |
| Often | 35 | 1.03 (0.73, 1.45) | 0.887 | 0.97 (0.61-1.53) | 0.881 |
| Per category increase | 1,076 | 0.93 (0.82, 1.05) | 0.253 | 0.85 (0.73, 1.00) | 0.054 |
| **Daytime napping** |  |  |  |  |  |
| Never/rarely | 606 | Reference |  | Reference |  |
| Sometimes | 425 | 1.00 (0.89, 1.14) | 0.939 | 0.96 (0.82, 1.13) | 0.657 |
| Usually | 48 | 1.01 (0.75, 1.36) | 0.957 | 0.97 (0.66, 1.44) | 0.892 |
| Per category increase | 1,079 | 1.00 (0.90, 1.12) | 0.931 | 0.97 (0.85, 1.11) | 0.685 |
| **Snoring** | |  |  |  |  |
| No | 642 | Reference |  | Reference |  |
| Yes | 345 | 1.01 (0.89, 1.16) | 0.849 | 1.03 (0.87, 1.22) | 0.705 |
| * |  | 1.02 (0.89, 1.17) | 0.763 | 1.03 (0.87, 1.21) | 0.768 |
| ** |  | 1.01 (0.89, 1.16) | 0.859 | 1.03 (0.87, 1.22) | 0.711 |
| *** |  | 1.02 (0.89, 1.16) | 0.827 | 1.03 (0.87, 1.22) | 0.705 |
| **Ovarian cancer** | | | | | |
| **Sleep duration** | |  |  |  |  |
| <7 h | 165 | 1.09 (0.91, 1.31) | 0.333 | 1.16 (0.93, 1.44) | 0.180 |
| 7-8 h | 447 | Reference |  | Reference |  |
| >8 h | 62 | 1.19 (0.91, 1.56) | 0.199 | 1.22 (0.88, 1.69) | 0.238 |
| Per 1h |  | 1.00 (0.93, 1.07) | 0.967 | 0.97 (0.89, 1.05) | 0.438 |
| **Chronotype** | |  |  |  |  |
| Morning | 380 | Reference |  | Reference |  |
| Evening | 228 | 1.06 (0.90, 1.26) | 0.457 | 1.07 (0.87, 1.30) | 0.519 |
| **Chronotype** |  |  |  |  |  |
| Definitely morning | 162 | Reference |  | Reference |  |
| More morning | 218 | 1.00 (0.82, 1.23) | 0.974 | 0.97 (0.76, 1.23) | 0.775 |
| More evening | 165 | 1.00 (0.81, 1.25) | 0.984 | 0.97 (0.75, 1.26) | 0.810 |
| Definitely evening | 63 | 1.27 (0.95, 1.71) | 0.107 | 1.31 (0.93, 1.85) | 0.126 |
| Mixed | 61 | 1.15 (0.86, 1.55) | 0.349 | 0.97 (0.65, 1.45) | 0.893 |
| **Insomnia** | |  |  |  |  |
| Never/rarely | 144 | Reference |  | Reference |  |
| Sometimes | 341 | 0.85 (0.70, 1.04) | 0.110 | 1.02 (0.80, 1.31) | 0.863 |
| Usually | 194 | 0.77 (0.62, 0.96) | 0.021 | 0.94 (0.71, 1.23) | 0.639 |
| No (ref) vs Yes | 535 | 0.85 (0.70, 1.04) | 0.110 | 0.99 (0.78, 1.26) | 0.958 |
| **Daytime sleepiness** |  |  |  |  |  |
| Never/rarely | 512 | Reference |  | Reference |  |
| Sometimes | 151 | 1.16 (0.96, 1.39) | 0.115 | 1.37 (1.10, 1.71) | 0.004 |
| Often | 14 | 0.92 (0.54-1.57) | 0.766 | 0.99 (0.51, 1.91) | 0.965 |
| Per category increase | 677 | 1.09 (0.93, 1.27) | 0.285 | 1.23 (1.03, 1.47) | 0.026 |
| **Daytime napping** |  |  |  |  |  |
| Never/rarely | 418 | Reference |  | Reference |  |
| Sometimes | 235 | 0.97 (0.82, 1.14) | 0.678 | 1.07 (0.88, 1.30) | 0.507 |
| Usually | 27 | 1.14 (0.77, 1.70) | 0.515 | 1.06 (0.63, 1.78) | 0.835 |
| Per category increase | 680 | 1.00 (0.87, 1.15) | 0.979 | 1.05 (0.89, 1.28) | 0.532 |
| **Snoring** | |  |  |  |  |
| No | 457 | Reference |  | Reference |  |
| Yes | 152 | 0.82 (0.68, 1.00) | 0.044 | 0.78 (0.62, 0.98) | 0.035 |
| * |  | 0.82 (0.68, 1.00) | 0.044 | 0.79 (0.63, 0.99) | 0.043 |
| ** |  | 0.82 (0.68, 0.99) | 0.038 | 0.77 (0.61, 0.97) | 0.029 |
| *** |  | 0.83 (0.68, 1.00) | 0.044 | 0.78 (0.62, 0.99) | 0.038 |

^a^ Adjusted for age, BMI, Townsend deprivation index.

^b^ Additionally adjusted for smoking status, coffee intake, physical activity (MET h/wk), education, tea intake, and history of PSA testing.

^c^ Additionally adjusted for smoking status, coffee intake, physical activity (MET h/wk), education, tea intake,

menopause status, hormone replacement therapy, and parity.

*Further adjusted for sleep duration (h/d).

**Further adjusted for insomnia.

*** Further adjusted for sleep apnoea.

**Table S6.** Associations of sleep traits with prostate cancer incidence in the UK Biobank population stratified by median age.

|  | **Events** | **Adjusted HR^a^ (95% CI)** | **P-value** | **Adjusted HR^b^**  **(95% CI)** | **P-value** |
| --- | --- | --- | --- | --- | --- |
| **Prostate cancer** | | | | | |
| **Age > 58** | | | | | |
| **Sleep duration** | |  |  |  |  |
| <7 h | 1,284 | 0.97 (0.91, 1.03) | 0.319 | 0.98 (0.91, 1.05) | 0.570 |
| 7-8 h | 4,311 | Reference |  | Reference |  |
| >8 h | 573 | 0.93 (0.85, 1.01) | 0.099 | 0.90 (0.81, 1.00) | 0.042 |
| Per 1h |  | 1.00 (0.98, 1.02) | 0.914 | 0.96 (0.97, 1.01) | 0.333 |
| **Chronotype** | |  |  |  |  |
| Morning | 3,542 | Reference |  | Reference |  |
| Evening | 1,776 | 0.97 (0.92, 1.03) | 0.358 | 0.96 (0.90, 1.03) | 0.273 |
| **Insomnia** | |  |  |  |  |
| Never/rarely | 1,697 | Reference |  | Reference |  |
| Sometimes | 2,855 | 1.06 (1.00, 1.12) | 0.074 | 1.07 (1.00, 1.15) | 0.065 |
| Usually | 1,638 | 1.07 (1.00, 1.15) | 0.044 | 1.06 (0.98, 1.15) | 0.178 |
| No (ref) vs Yes | 6,190 | 1.06 (1.00, 1.12) | 0.035 | 1.06 (1.00, 1.14) | 0.623 |
| **Daytime sleepiness** |  |  |  |  |  |
| Never/rarely | 4,315 | Reference |  | Reference |  |
| Sometimes | 1,639 | 1.01 (0.95, 1.07) | 0.723 | 1.00 (0.94-1.07) | 0.928 |
| Often | 216 | 1.04 (0.91-1.20) | 0.550 | 1.03 (0.94-1.07) | 0.723 |
| Per category increase | 6,170 | 1.01 (0.97, 1.06) | 0.540 | 1.01 (0.95, 1.07) | 0.786 |
| **Daytime napping** |  |  |  |  |  |
| Never/rarely | 2,673 | Reference |  | Reference |  |
| Sometimes | 2,924 | 0.98 (0.93, 1.03) | 0.405 | 0.99 (0.93, 1.05) | 0.640 |
| Usually | 596 | 0.94 (0.86, 1.03) | 0.210 | 1.00 (0.90, 1.1) | 0.985 |
| Per category increase | 6,193 | 0.97 (0.94, 1.01) | 0.188 | 0.99 (0.95, 1.04) | 0.803 |
| **Snoring** | |  |  |  |  |
| No | 3,026 | Reference |  | Reference |  |
| Yes | 2,840 | 1.05 (1.00, 1.11) | 0.070 | 1.04 (0.98, 1.11) | 0.210 |
| * |  | 1.05 (1.00, 1.11) | 0.068 | 1.04 (0.98, 1.11) | 0.179 |
| ** |  | 1.05 (1.00, 1.11) | 0.062 | 1.04 (0.98, 1.11) | 0.192 |
| *** |  | 1.05 (1.00, 1.11) | 0.067 | 1.04 (0.98, 1.11) | 0.201 |
| **Age ≤ 58** | | | | | |
| **Sleep duration** |  |  |  |  |  |
| <7 h | 637 | 0.98 (0.90, 1.08) | 0.693 | 0.97 (0.87, 1.08) | 0.610 |
| 7-8 h | 1,657 | Reference |  | Reference |  |
| >8 h | 112 | 1.03 (0.83, 1.28) | 0.796 | 1.07 (0.86, 1.33) | 0.552 |
| Per 1h |  | 1.02 (0.98,1.06) | 0.429 | 1.02 (0.98,1.07) | 0.352 |
| **Chronotype** |  |  |  |  |  |
| Morning | 1,234 | Reference |  | Reference |  |
| Evening | 859 | 1.07 (0.98, 1.17) | 0.124 | 1.14 (1.03, 1.26) | 0.012 |
| **Insomnia** |  |  |  |  |  |
| Never/rarely | 745 | Reference |  | Reference |  |
| Sometimes | 1,137 | 1.05 (0.96, 1.15) | 0.324 | 1.01 (0.91, 1.13) | 0.821 |
| Usually | 529 | 1.04 (0.93, 1.16) | 0.493 | 0.98 (0.86, 1.12) | 0.760 |
| No (ref) vs Yes | 2,411 | 1.05 (0.96, 1.14) | 0.317 | 1.00 (0.91, 1.11) | 0.966 |
| **Daytime sleepiness** |  |  |  |  |  |
| Never/rarely | 1,895 | Reference |  | Reference |  |
| Sometimes | 460 | 1.01 (0.69, 1.07) | 0.723 | 0.74 (0.52, 1.05) | 0.921 |
| Often | 52 | 0.91 (0.91, 1.20) | 0.550 | 0.98 (0.86, 1.10) | 0.684 |
| Per category increase | 2,407 | 0.99 (0.91, 1.08) | 0.801 | 0.94 (0.85, 1.04) | 0.198 |
| **Daytime napping** |  |  |  |  |  |
| Never/rarely | 1,387 | Reference |  | Reference |  |
| Sometimes | 897 | 0.98 (0.90, 1.06) | 0.604 | 0.95 (0.86, 1.05) | 0.289 |
| Usually | 126 | 0.95 (0.79, 1.15) | 0.619 | 1.08 (0.88, 1.32) | 0.478 |
| Per category increase | 2,410 | 0.98 (0.91, 1.05) | 0.511 | 0.99 (0.91, 1.07) | 0.775 |
| **Snoring** |  |  |  |  |  |
| No | 1,156 | Reference |  | Reference |  |
| Yes | 1,146 | 1.01 (0.93, 1.10) | 0.823 | 0.97 (0.88, 1.07) | 0.562 |
| * |  | 1.01 (0.93, 1.10) | 0.795 | 0.97 (0.88, 1.07) | 0.564 |
| ** |  | 1.01 (0.93, 1.10) | 0.845 | 0.97 (0.88, 1.07) | 0.523 |
| *** |  | 1.01 (0.93, 1.10) | 0.772 | 0.98 (0.89, 1.07) | 0.616 |

^a^ Adjusted for Age, BMI, Townsend deprivation index

^b^ Additionally adjusted for Smoking status, Coffee intake, Physical activity (MET/h-wk), Education, Tea intake, History of PSA testing

*Further adjusted for sleep duration (h/d)

**Further adjusted for insomnia

*** Further adjusted for sleep apnoea

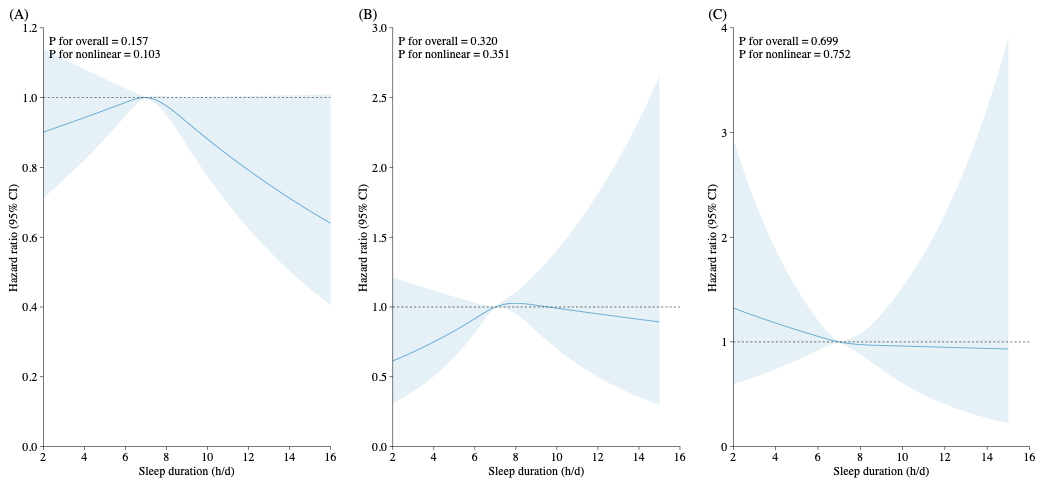

**Figure S4**. Non-linear relationship between sleep duration and (A) prostate, (B) endometrial, and (C) epithelial ovarian cancers using restricted cubic splines. Knots were placed at the 5th, 50th, and 95th percentile, with the 50th (median) as the reference.

**Table S7**. Associations of sleep traits with endometrial cancer incidence in the UK Biobank population stratified by median age.

|  | **Events** | **Adjusted HR^a^ (95% CI)** | **P-value** | **Adjusted HR^b^ (95% CI)** | **P-value** |
| --- | --- | --- | --- | --- | --- |
| **Endometrial cancer** | | | | | |
| **Age >56** | | | | | |
| **Sleep duration** | |  |  |  |  |
| <7 h | 168 | 0.94 (0.79, 1.12) | 0.503 | 0.91 (0.73, 1.13) | 0.393 |
| 7-8 h | 461 | Reference |  | Reference |  |
| >8 h | 74 | 1.09 (0.85, 1.39) | 0.511 | 0.97 (0.71, 1.34) | 0.876 |
| Per 1h |  | 1.07 (1.00, 1.14) | 0.051 | 1.03 (0.96, 1.12) | 0.402 |
| **Chronotype** | |  |  |  |  |
| Morning | 424 | Reference |  | Reference |  |
| Evening | 233 | 0.99 (0.84, 1.16) | 0.909 | 0.99 (0.81, 1.21) | 0.955 |
| **Insomnia** | |  |  |  |  |
| Never/rarely | 110 | Reference |  | Reference |  |
| Sometimes | 371 | 1.04 (0.84, 1.28) | 0.729 | 1.16 (0.89, 1.51) | 0.286 |
| Usually | 231 | 0.91 (0.73, 1.15) | 0.434 | 1.04 (0.78, 1.38) | 0.790 |
| No (ref) vs Yes | 712 | 0.99 (0.81, 1.21) | 0.896 | 1.11 (0.86, 1.43) | 0.427 |
| **Daytime sleepiness** |  |  |  |  |  |
| Never/rarely | 528 | Reference |  | Reference |  |
| Sometimes | 158 | 0.86 (0.72, 1.03) | 0.10 | 0.75 (0.59, 0.94) | 0.014 |
| Often | 25 | 1.08 (0.72, 1.62) | 0.705 | 0.89 (0.51, 1.56) | 0.685 |
| Per category increase | 711 | 0.93 (0.80, 1.07) | 0.313 | 0.81 (0.67, 0.99) | 0.035 |
| **Daytime napping** |  |  |  |  |  |
| Never/rarely | 383 | Reference |  | Reference |  |
| Sometimes | 291 | 0.99 (0.85, 1.16) | 0.929 | 0.95 (0.78, 1.15) | 0.610 |
| Usually | 38 | 1.15 (0.82, 1.61) | 0.427 | 1.09 (0.71, 1.68) | 0.697 |
| Per category increase | 699 | 1.03 (0.90, 1.17) | 0.696 | 0.99 (0.84, 1.16) | 0.884 |
| **Snoring** | |  |  |  |  |
| No | 439 | Reference |  | Reference |  |
| Yes | 218 | 0.93 (0.79, 1.10) | 0.403 | 0.92 (0.75, 1.13) | 0.445 |
| * |  | 0.93 (0.79, 1.10) | 0.421 | 0.91 (0.74, 1.12) | 0.383 |
| ** |  | 0.93 (0.79, 1.10) | 0.392 | 0.92 (0.75, 1.13) | 0.437 |
| *** |  | 0.93 (0.79, 1.10) | 0.411 | 0.92 (0.75, 1.13) | 0.411 |
| **Age ≤ 56** | | | | | |
| **Sleep duration** |  |  |  |  |  |
| <7 h | 90 | 0.97 (0.76, 1.23) | 0.777 | 0.83 (0.60, 1.16) | 0.279 |
| 7-8 h | 249 | Reference |  | Reference |  |
| >8 h | 25 | 0.94 (0.62, 1.42) | 0.765 | 0.85 (0.49, 1.47) | 0.564 |
| Per 1h |  | 1.03 (0.94,1.13) | 0.535 | 1.07 (0.95, 1.21) | 0.274 |
| **Chronotype** |  |  |  |  |  |
| Morning | 181 | Reference |  | Reference |  |
| Evening | 151 | 1.21 (0.97, 1.51) | 0.084 | 1.25 (0.95, 1.65) | 0.106 |
| **Insomnia** |  |  |  |  |  |
| Never/rarely | 83 | Reference |  | Reference |  |
| Sometimes | 173 | 0.88 (0.68, 1.14) | 0.340 | 0.92 (0.66, 1.28) | 0.625 |
| Usually | 111 | 0.79 (0.59, 1.05) | 0.102 | 0.99 (0.69, 1.43) | 0.949 |
| No (ref) vs Yes | 367 | 0.84 (0.66, 1.08) | 0.173 | 0.94 (0.69, 1.29) | 0.722 |
| **Daytime sleepiness** |  |  |  |  |  |
| Never/rarely | 294 | Reference |  | Reference |  |
| Sometimes | 61 | 0.95 (0.72, 1.26) | 0.720 | 0.90 (0.62, 1.31) | 0.587 |
| Often | 10 | 0.91 (0.47, 1.76) | 0.772 | 1.19 (0.53, 2.70) | 0.674 |
| Per category increase | 365 | 0.95 (0.76, 1.19) | 0.657 | 0.98 (0.73, 1.31) | 0.872 |
| **Daytime napping** |  |  |  |  |  |
| Never/rarely | 223 | Reference |  | Reference |  |
| Sometimes | 134 | 1.05 (0.85, 1.31) | 0.648 | 1.01 (0.77, 1.34) | 0.923 |
| Usually | 10 | 0.67 (0.33, 1.36) | 0.267 | 0.62 (0.23, 1.69) | 0.354 |
| Per category increase | 367 | 0.98 (0.81, 1.18) | 0.822 | 0.95 (0.74, 1.22) | 0.697 |
| **Snoring** |  |  |  |  |  |
| No | 203 | Reference |  | Reference |  |
| Yes | 127 | 1.14 (0.90, 1.43) | 0.278 | 1.27 (0.95, 1.70) | 0.110 |
| * |  | 1.15 (0.91, 1.45) | 0.231 | 1.27 (0.95, 1.71) | 0.108 |
| ** |  | 1.14 (0.90, 1.43) | 0.278 | 1.27 (0.95, 1.70) | 0.109 |
| *** |  | 1.14 (0.91, 1.44) | 0.265 | 1.27 (0.95, 1.71) | 0.103 |

^a^ Adjusted for Age, BMI, Townsend deprivation index

^b^Additionally adjusted for Smoking status, Coffee intake, Physical activity (MET/h-wk), Education, Tea intake,

Menopause status, Hormone replacement therapy, Parity

*Further adjusted for sleep duration (h/d)

**Further adjusted for insomnia

*** Further adjusted for sleep apnoea

**Table S8.** Associations of sleep traits with epithelial ovarian cancer incidence in the UK Biobank stratified by median age.

|  | **Events** | **Adjusted HR^a^ (95% CI)** | **P-value** | **Adjusted HR^b^ (95% CI)** | **P-value** |
| --- | --- | --- | --- | --- | --- |
| **Ovarian cancer** | | | | | |
| **Age >56** | | | | | |
| **Sleep duration** | |  |  |  |  |
| <7 h | 105 | 1.08 (0.86, 1.36) | 0.481 | 1.11 (0.84, 1.48) | 0.458 |
| 7-8 h | 270 | Reference |  | Reference |  |
| >8 h | 41 | 1.15 (0.83, 1.61) | 0.397 | 1.28 (0.86, 1.91) | 0.225 |
| Per 1h |  | 1.02 (0.94, 1.12) | 0.620 | 1.00 (0.90, 1.12) | 0.999 |
| **Chronotype** | |  |  |  |  |
| Morning | 242 | Reference |  | Reference |  |
| Evening | 136 | 1.09 (0.89, 1.35) | 0.400 | 1.18 (0.91, 1.53) | 0.221 |
| **Insomnia** | |  |  |  |  |
| Never/rarely | 80 | Reference |  | Reference |  |
| Sometimes | 219 | 0.82 (0.63, 1.06) | 0.124 | 1.13 (0.80, 1.60) | 0.477 |
| Usually | 122 | 0.72 (0.54, 0.95) | 0.020 | 0.94 (0.64, 1.37) | 0.734 |
| No (ref) vs Yes | 421 | 0.78 (0.61, 0.99) | 0.432 | 1.06 (0.76, 1.48) | 0.736 |
| **Daytime sleepiness** |  |  |  |  |  |
| Never/rarely | 306 | Reference |  | Reference |  |
| Sometimes | 105 | 1.12(0.90, 1.41) | 0.303 | 1.39 (1.06, 1.82) | 0.017 |
| Often | 8 | 0.79 (0.39, 1.60) | 0.519 | 1.03 (0.45, 2.32) | 0.950 |
| Per category increase | 419 | 1.05 (0.87, 1.26) | 0.643 | 1.25 (1.00, 1.56) | 0.534 |
| **Daytime napping** |  |  |  |  |  |
| Never/rarely | 255 | Reference |  | Reference |  |
| Sometimes | 146 | 0.88 (0.72, 1.08) | 0.228 | 0.92 (0.71, 1.19) | 0.533 |
| Usually | 20 | 1.22 (0.77, 1.92) | 0.401 | 1.17 (0.65, 2.11) | 0.602 |
| Per category increase | 421 | 0.96 (0.81, 1.14) | 0.661 | 0.98 (0.79, 1.21) | 0.854 |
| **Snoring** | |  |  |  |  |
| No | 274 | Reference |  | Reference |  |
| Yes | 98 | 0.87 (0.69, 1.10) | 0.245 | 0.75 (0.56, 1.02) | 0.066 |
| * |  | 0.87 (0.69, 1.10) | 0.241 | 0.76 (0.56, 1.03) | 0.075 |
| ** |  | 0.87 (0.69, 1.10) | 0.239 | 0.75 (0.56, 1.02) | 0.064 |
| *** |  | 0.87 (0.69, 1.10) | 0.237 | 0.76 (0.56, 1.02) | 0.070 |
| **Age ≤56** | | | | | |
| **Sleep duration** |  |  |  |  |  |
| <7 h | 60 | 1.10 (0.82, 1.48) | 0.535 | 1.21 (0.86, 1.71) | 0.266 |
| 7-8 h | 177 | Reference |  | Reference |  |
| >8 h | 21 | 1.27 (0.81, 2.00) | 0.304 | 1.13 (0.64, 2.00) | 0.679 |
| Per 1h |  | 0.97 (0.86, 1.09) | 0.588 | 0.92 (0.80, 1.07) | 0.279 |
| **Chronotype** |  |  |  |  |  |
| Morning | 138 | Reference |  | Reference |  |
| Evening | 92 | 1.01 (0.78, 1.32) | 0.919 | 0.94 (0.69, 1.28) | 0.679 |
| **Insomnia** |  |  |  |  |  |
| Never/rarely | 64 | Reference |  | Reference |  |
| Sometimes | 122 | 0.89 (0.65, 1.20) | 0.435 | 0.89 (0.62, 1.27) | 0.514 |
| Usually | 72 | 0.86 (0.61, 1.21) | 0.380 | 0.95 (0.64, 1.42) | 0.805 |
| No (ref) vs Yes | 258 | 0.88 (0.66, 1.17) | 0.363 | 0.91 (0.65, 1.28) | 0.591 |
| **Daytime sleepiness** |  |  |  |  |  |
| Never/rarely | 206 | Reference |  | Reference |  |
| Sometimes | 46 | 1.24 (0.90, 1.71) | 0.197 | 1.36 (0.94, 1.97) | 0.106 |
| Often | 6 | 1.17 (0.52, 2.65) | 0.698 | 0.93 (0.30, 2.92) | 0.901 |
| Per category increase | 258 | 1.17 (0.91, 1.52) | 0.225 | 1.21 (0.89, 1.64) | 0.230 |
| **Daytime napping** |  |  |  |  |  |
| Never/rarely | 163 | Reference |  | Reference |  |
| Sometimes | 89 | 1.12 (0.87, 1.46) | 0.382 | 1.32 (0.98, 1.78) | 0.07 |
| Usually | 7 | 0.91 (0.40, 2.05) | 0.815 | 0.73 (0.23, 2.32) | 0.599 |
| Per category increase | 259 | 1.07 (0.85, 1.34) | 0.555 | 1.18 (0.91, 1.54) | 0.212 |
| **Snoring** |  |  |  |  |  |
| No | 183 | Reference |  | Reference |  |
| Yes | 54 | 0.74 (0.54, 1.01) | 0.060 | 0.80 (0.56, 1.15) | 0.229 |
| * |  | 0.74 (0.54, 1.02) | 0.063 | 0.80 (0.56, 1.16) | 0.247 |
| ** |  | 0.73 (0.53, 1.00) | 0.047 | 0.78 (0.54, 1.13) | 0.188 |
| *** |  | 0.74 (0.54, 1.02) | 0.062 | 0.80 (0.56, 1.15) | 0.235 |

^a^ Adjusted for Age, BMI, Townsend deprivation index

^b^Additionally adjusted for Smoking status, Coffee intake, Physical activity (MET/h-wk), Education, Tea intake,

Menopause status, Hormone replacement therapy, Parity

*Further adjusted for sleep duration (h/d)

**Further adjusted for insomnia

*** Further adjusted for sleep apnoea.

**Table S9**. Associations of sleep traits with prostate cancer incidence in the UK Biobank stratified by median body mass index.

|  | **Events** | **Adjusted HR^a^**  **(95% CI)** | **P-value** | **Adjusted HR^b^**  **(95% CI)** | **P-value** |
| --- | --- | --- | --- | --- | --- |
| **Prostate cancer** | | | | | |
| **BMI >27** | | | | | |
| **Sleep duration** | |  |  |  |  |
| <7 h | 1,070 | 0.99 (0.92, 1.06) | 0.793 | 1.01 (0.93, 1.10) | 0.829 |
| 7-8 h | 2,967 | Reference |  | Reference |  |
| >8 h | 404 | 0.97 (0.88, 1.08) | 0.621 | 0.96 (0.85, 1.09) | 0.520 |
| Per 1h |  | 1.00 (0.97, 1.03) | 0.917 | 0.99 (0.95, 1.02) | 0.368 |
| **Chronotype** | |  |  |  |  |
| Morning | 2,496 | Reference |  | Reference |  |
| Evening | 1,362 | 0.98 (0.92, 1.05) | 0.544 | 0.98 (0.91, 1.06) | 0.680 |
| **Insomnia** | |  |  |  |  |
| Never/rarely | 1,220 | Reference |  | Reference |  |
| Sometimes | 2,071 | 1.06 (0.99, 1.14) | 0.095 | 1.06 (0.98, 1.15) | 0.171 |
| Usually | 1,167 | 1.06 (0.98, 1.15) | 0.015 | 1.02 (0.93, 1.12) | 0.681 |
| No (ref) vs Yes | 4,458 | 1.06 (0.99, 1.13) | 0.075 | 1.05 (0.97, 1.13) | 0.257 |
| **Daytime sleepiness** |  |  |  |  |  |
| Never/rarely | 3,151 | Reference |  | Reference |  |
| Sometimes | 1,149 | 1.01 (0.94, 1.08) | 0.738 | 1.01 (0.93, 1.09) | 0.862 |
| Often | 146 | 0.93 (0.79, 1.10) | 0.382 | 0.89 (0.72, 1.09) | 0.251 |
| Per category increase | 4,446 | 0.99 (0.94, 1.05) | 0.784 | 0.98 (0.92, 1.05) | 0.587 |
| **Daytime napping** |  |  |  |  |  |
| Never/rarely | 1,929 | Reference |  | Reference |  |
| Sometimes | 2,137 | 0.99 (0.93, 1.06) | 0.816 | 0.99 (0.92, 1.07) | 0.851 |
| Usually | 393 | 0.88 (0.79, 0.99) | 0.026 | 0.96 (0.84, 1.09) | 0.488 |
| Per category increase | 4,459 | 0.96 (0.92, 1.01) | 0.089 | 0.98 (0.93, 1.04) | 0.557 |
| **Snoring** | |  |  |  |  |
| No | 1,877 | Reference |  | Reference |  |
| Yes | 2,364 | 1.07 (1.01, 1.14) | 0.024 | 1.05 (0.98, 1.13) | 0.201 |
| * |  | 1.07 (1.01, 1.14) | 0.027 | 1.05 (0.98, 1.13) | 0.187 |
| ** |  | 1.07 (1.01, 1.14) | 0.016 | 1.05 (0.98, 1.13) | 0.200 |
| *** |  | 1.07 (1.01, 1.14) | 0.021 | 1.05 (0.98, 1.13) | 0.184 |
| **BMI ≤ 27** | | | | | |
| **Sleep duration** |  |  |  |  |  |
| <7 h | 842 | 0.97 (0.90, 1.05) | 0.477 | 0.96 (0.88, 1.06) | 0.426 |
| 7-8 h | 2,985 | Reference |  | Reference |  |
| >8 h | 281 | 0.86 (0.76, 0.98) | 0.019 | 0.84 (0.73, 0.98) | 0.024 |
| Per 1h |  | 1.00 (0.97, 1.03) | 0.763 | 0.99 (0.95, 1.03) | 0.553 |
| **Chronotype** |  |  |  |  |  |
| Morning | 2,265 | Reference |  | Reference |  |
| Evening | 1,267 | 1.01 (0.95, 1.09) | 0.717 | 1.03 (0.95, 1.12) | 0.482 |
| **Insomnia** |  |  |  |  |  |
| Never/rarely | 1,219 | Reference |  | Reference |  |
| Sometimes | 1,910 | 1.04 (0.97, 1.12) | 0.239 | 1.04 (0.96, 1.13) | 0.311 |
| Usually | 989 | 1.06 (0.98, 1.12) | 0.143 | 1.04 (0.95, 1.15) | 0.389 |
| No (ref) vs Yes | 4,118 | 1.05 (0.98, 1.12) | 0.145 | 1.04 (0.97, 1.13) | 0.277 |
| **Daytime sleepiness** |  |  |  |  |  |
| Never/rarely | 3,044 | Reference |  | Reference |  |
| Sometimes | 944 | 0.99 (0.92, 1.06) | 0.698 | 0.96 (0.88, 1.05) | 0.352 |
| Often | 119 | 1.14 (0.95, 1.37) | 0.172 | 1.06 (0.85, 1.33) | 0.579 |
| Per category increase | 4,107 | 1.01 (0.95, 1.08) | 0.649 | 0.98 (0.92, 1.06) | 0.674 |
| **Daytime napping** |  |  |  |  |  |
| Never/rarely | 2,123 | Reference |  | Reference |  |
| Sometimes | 1,671 | 0.95 (0.89, 1.01) | 0.113 | 0.94 (0.87, 1.02) | 0.129 |
| Usually | 325 | 0.99 (0.88, 1.12) | 0.923 | 1.04 (0.91, 1.19) | 0.568 |
| Per category increase | 4,119 | 0.98 (0.93, 1.02) | 0.321 | 0.99 (0.93, 1.04) | 0.618 |
| **Snoring** |  |  |  |  |  |
| No | 2,295 | Reference |  | Reference |  |
| Yes | 1,607 | 1.02 (0.96, 1. 09) | 0.467 | 1.01 (0.94, 1.09) | 0.703 |
| * |  | 1.03 (0.96, 1.10) | 0.400 | 1.02 (0.94, 1.10) | 0.652 |
| ** |  | 1.02 (0.96, 1.09) | 0.466 | 1.02 (0.94, 1.09) | 0.696 |
| *** |  | 1.02 (0.96, 1.09) | 0.458 | 1.02 (0.94, 1.09) | 0.674 |

^a^ Adjusted for Age, BMI, Townsend deprivation index

^b^ Additionally adjusted for Smoking status, Coffee intake, Physical activity (MET/h-wk), Education, Tea intake, History of PSA testing

*Further adjusted for sleep duration (h/d)

**Further adjusted for insomnia

*** Further adjusted for sleep apnoea

**Table S10**. Associations of sleep traits with endometrial cancer incidence in the UK Biobank stratified by median body mass index.

|  | **Events** | **Adjusted HR^a^ (95% CI)** | **P-value** | **Adjusted HR^b^**  **(95% CI)** | **P-value** |
| --- | --- | --- | --- | --- | --- |
| **Endometrial cancer** | | | | | |
| **BMI >26** | | | | | |
| **Sleep duration** | |  |  |  |  |
| <7 h | 189 | 0.92 (0.78, 1.09) | 0.332 | 0.87 (0.70, 1.08) | 0.220 |
| 7-8 h | 496 | Reference |  | Reference |  |
| >8 h | 81 | 1.06 (0.84, 1.34) | 0.636 | 0.88 (0.63, 1.21) | 0.426 |
| Per 1h |  | 1.05 (0.99, 1.12) | 0.087 | 1.03 (0.96, 1.11) | 0.431 |
| **Chronotype** | |  |  |  |  |
| Morning | 428 | Reference |  | Reference |  |
| Evening | 281 | 1.04 (0.89, 1.21) | 0.641 | 1.06 (0.87, 1.28) | 0.566 |
| **Insomnia** | |  |  |  |  |
| Never/rarely | 140 | Reference |  | Reference |  |
| Sometimes | 383 | 0.95 (0.78, 1.15) | 0.582 | 1.06 (0.83, 1.35) | 0.666 |
| Usually | 251 | 0.82 (0.67, 1.01) | 0.061 | 0.91 (0.70, 1.19) | 0.495 |
| No (ref) vs Yes | 774 | 0.89 (0.74, 1.07) | 0.225 | 1.00 (0.79, 1.26) | 0.964 |
| **Daytime sleepiness** |  |  |  |  |  |
| Never/rarely | 579 | Reference |  | Reference |  |
| Sometimes | 169 | 0.87 (0.73, 1.03) | 0.112 | 0.79 (0.62, 0.99) | 0.396 |
| Often | 25 | 0.90 (0.60, 1.35) | 0.608 | 0.83 (0.48, 1.45) | 0.510 |
| Per category increase | 773 | 0.90 (0.78, 1.04) | 0.140 | 0.83 (0.69, 1.00) | 0.053 |
| **Daytime napping** |  |  |  |  |  |
| Never/rarely | 416 | Reference |  | Reference |  |
| Sometimes | 322 | 0.97 (0.83, 1.12) | 0.655 | 0.95 (0.78, 1.14) | 0.562 |
| Usually | 36 | 0.93 (0.66, 1.31) | 0.681 | 0.98 (0.63, 1.53) | 0.939 |
| Per category increase | 774 | 0.97 (0.85, 1.09) | 0.580 | 0.96 (0.82, 1.13) | 0.645 |
| **Snoring** | |  |  |  |  |
| No | 415 | Reference |  | Reference |  |
| Yes | 288 | 1.06 (0.91, 1.23) | 0.464 | 1.07 (0.88, 1.30) | 0.480 |
| * |  | 1.06 (0.91, 1.24) | 0.438 | 1.06 (0.87, 1.28) | 0.554 |
| ** |  | 1.06 (0.91, 1.23) | 0.477 | 1.07 (0.88, 1.30) | 0.492 |
| *** |  | 1.06 (0.91, 1.23) | 0.446 | 1.07 (0.88, 1.30) | 0.480 |
| **BMI ≤26** | | | | | |
| **Sleep duration** |  |  |  |  |  |
| <7 h | 68 | 1.07 (0.81, 1.41) | 0.621 | 0.92 (0.65, 1.30) | 0.650 |
| 7-8 h | 209 | Reference |  | Reference |  |
| >8 h | 18 | 0.93 (0.57, 1.50) | 0.752 | 1.13 (0.67, 1.89) | 0.651 |
| Per 1h |  | 1.03 (0.92, 1.16) | 0.575 | 1.08 (0.95, 1.24) | 0.252 |
| **Chronotype** |  |  |  |  |  |
| Morning | 173 | Reference |  | Reference |  |
| Evening | 101 | 1.12 (0.88, 1.43) | 0.369 | 1.09 (0.81, 1.46) | 0.583 |
| **Insomnia** |  |  |  |  |  |
| Never/rarely | 53 | Reference |  | Reference |  |
| Sometimes | 157 | 1.10 (0.80, 1.50) | 0.561 | 1.12 (0.77, 1.64) | 0.551 |
| Usually | 89 | 1.08 (0.77, 1.53) | 0.643 | 1.35 (0.90, 2.02) | 0.151 |
| No (ref) vs Yes | 299 | 1.09 (0.81, 1.47) | 0.516 | 1.20 (0.84, 1.73) | 0.311 |
| **Daytime sleepiness** |  |  |  |  |  |
| Never/rarely | 239 | Reference |  | Reference |  |
| Sometimes | 49 | 0.91 (0.61, 1.24) | 0.546 | 0.77 (0.52, 1.14) | 0.191 |
| Often | 9 | 1.57 (0.81, 3.06) | 1.83 | 1.48 (0.65, 3.34) | 0.349 |
| Per category increase | 297 | 1.03 (0.81, 1.32) | 0.794 | 0.92 (0.67, 1.26) | 0.601 |
| **Daytime napping** |  |  |  |  |  |
| Never/rarely | 187 | Reference |  | Reference |  |
| Sometimes | 102 | 1.11 (0.87, 1.42) | 0.391 | 1.01 (0.76, 1.36) | 0.930 |
| Usually | 10 | 1.31 (0.69, 2.48) | 0.405 | 0.90 (0.37, 2.20) | 0.814 |
| Per category increase | 299 | 1.12 (0.91, 1.38) | 0.270 | 0.99 (0.77, 1.28) | 0.963 |
| **Snoring** |  |  |  |  |  |
| No | 224 | Reference |  | Reference |  |
| Yes | 55 | 0.89 (0.66, 1.20) | 0.436 | 0.95 (0.67, 1.35) | 0.769 |
| * |  | 0.91 (0.67, 1.22) | 0.519 | 0.95 (0.67, 1.35) | 0.789 |
| ** |  | 0.89 (0.66, 1.19) | 0.434 | 0.95 (0.67, 1.35) | 0.779 |
| *** |  | 0.89 (0.66, 1.20) | 0.442 | 0.95 (0.67, 1.35) | 0.776 |

^a^ Adjusted for Age, BMI, Townsend deprivation index

^b^Additionally adjusted for Smoking status, Coffee intake, Physical activity (MET/h-wk), Education, Tea intake,

Menopause status, Hormone replacement therapy, Parity

*Further adjusted for sleep duration (h/d)

**Further adjusted for insomnia

*** Further adjusted for sleep apnoea

**Table S11**. Associations of sleep traits with epithelial ovarian cancer incidence in the UK Biobank stratified by median body mass index.

|  | **Total (Events)** | **Adjusted HR^a^**  **(95% CI)** | **P-value** | **Adjusted HR^b^**  **(95% CI)** | **P-value** |
| --- | --- | --- | --- | --- | --- |
| **Ovarian cancer** | | | | | |
| **BMI >26** | | | | | |
| **Sleep duration** | |  |  |  |  |
| <7 h | 90 | 1.16 (0.90, 1.48) | 0.250 | 1.17 (0.86, 1.58) | 0.317 |
| 7-8 h | 209 | Reference |  | Reference |  |
| >8 h | 42 | 1.45 (1.04, 2.03) | 0.272 | 1.53 (1.02, 2.29) | 0.038 |
| Per 1h |  | 1.03 (0.93, 1.12) | 0.598 | 1.01 (0.90, 1.13) | 0.863 |
| **Chronotype** | |  |  |  |  |
| Morning | 187 | Reference |  | Reference |  |
| Evening | 118 | 1.03 (0.82, 1.30) | 0.783 | 1.12 (0.85, 1.48) | 0.405 |
| **Insomnia** | |  |  |  |  |
| Never/rarely | 65 | Reference |  | Reference |  |
| Sometimes | 170 | 0.92 (0.69, 1.22) | 0.549 | 0.99 (0.70, 1.41) | 0.965 |
| Usually | 109 | 0.85 (0.62, 1.16) | 0.302 | 0.89 (0.61, 1.31) | 0.560 |
| No (ref) vs Yes | 354 | 0.89 (0.68, 1.17) | 0.396 | 0.95 (0.68, 1.33) | 0.776 |
| **Daytime sleepiness** |  |  |  |  |  |
| Never/rarely | 254 | Reference |  | Reference |  |
| Sometimes | 82 | 1.11 (0.86, 1.42) | 0.431 | 1.36 (1.01, 1.83) | 0.446 |
| Often | 6 | 0.65 (0.29, 1.46) | 0.299 | 0.56 (0.18, 1.76) | 0.324 |
| Per category increase | 342 | 1.00 (0.81, 1.24) | 0.992 | 1.14 (0.89, 1.47) | 0.305 |
| **Daytime napping** |  |  |  |  |  |
| Never/rarely | 201 | Reference |  | Reference |  |
| Sometimes | 123 | 0.88 (0.70, 1.10) | 0.250 | 0.96 (0.70, 1.22) | 0.595 |
| Usually | 21 | 1.48 (0.94, 2.32) | 0.921 | 1.47 (0.83, 2.62) | 0.186 |
| Per category increase | 325 | 1.01 (0.84, 1.22) | 0.913 | 1.04 (0.83, 1.31) | 0.740 |
| **Snoring** | |  |  |  |  |
| No | 208 | Reference |  | Reference |  |
| Yes | 99 | 0.83 (0.65, 1.05) | 0.138 | 0.81 (0.60, 1.09) | 0.157 |
| * |  | 0.82 (0.65, 1.05) | 0.117 | 0.81 (0.60, 1.09) | 0.162 |
| ** |  | 0.82 (0.65, 1.05) | 0.119 | 0.80 (0.59, 1.07) | 0.132 |
| *** |  | 0.83 (0.65, 1.06) | 0.137 | 0.81 (0.61, 1.09) | 0.167 |
| **BMI ≤26** | | | | | |
| **Sleep duration** |  |  |  |  |  |
| <7 h | 74 | 1.03 (0.79, 1.34) | 0.832 | 1.15 (0.84, 1.58) | 0.371 |
| 7-8 h | 237 | Reference |  | Reference |  |
| >8 h | 19 | 0.87 (0.55, 1.39) | 0.566 | 0.83 (0.46, 1.50) | 0.541 |
| Per 1h |  | 0.97 (0.87, 1.08) | 0.618 | 0.91 (0.80, 1.04) | 0.178 |
| **Chronotype** |  |  |  |  |  |
| Morning | 191 | Reference |  | Reference |  |
| Evening | 109 | 1.10 (0.87, 1.39) | 0.426 | 1.02 (0.76, 1.36) | 0.903 |
| **Insomnia** |  |  |  |  |  |
| Never/rarely | 79 | Reference |  | Reference |  |
| Sometimes | 168 | 0.80 (0.61, 1.04) | 0.099 | 1.05 (0.74, 1.48) | 0.797 |
| Usually | 85 | 0.70 (0.52, 0.96) | 0.026 | 0.98 (0.67, 1.46) | 0.938 |
| No (ref) vs Yes | 332 | 0.76 (0.59, 0.98) | 0.038 | 1.03 (0.74, 1.43) | 0.867 |
| **Daytime sleepiness** |  |  |  |  |  |
| Never/rarely | 255 | Reference |  | Reference |  |
| Sometimes | 69 | 1.22 (0.94 1.60) | 0.142 | 1.39 (1.01, 1.91) | 0.044 |
| Often | 8 | 1.32 (0.65, 2.67) | 0.438 | 1.55 (0.69, 3.50) | 0.293 |
| Per category increase | 332 | 1.20 (0.96, 1.49) | 0.112 | 1.33 (1.03, 1.72) | 0.030 |
| **Daytime napping** |  |  |  |  |  |
| Never/rarely | 215 | Reference |  | Reference |  |
| Sometimes | 112 | 1.05 (0.86, 1.36) | 0.495 | 1.25 (0.95, 1.64) | 0.115 |
| Usually | 5 | 0.58 (0.24, 1.40) | 0.224 | 0.37 0.09, 1.48) | 0.158 |
| Per category increase | 333 | 0.99 (0.81, 1.22) | 0.953 | 1.07 (0.84, 1.37) | 0.564 |
| **Snoring** |  |  |  |  |  |
| No | 246 | Reference |  | Reference |  |
| Yes | 53 | 0.81 (0.60, 1.09) | 0.156 | 0.73 (0.50, 1.06) | 0.096 |
| * |  | 0.82 (0.61, 1.10) | 0.184 | 0.74 (0.51, 1.08) | 0.116 |
| ** |  | 0.81 (0.60, 1.09) | 0.158 | 0.73 (0.50, 1.06) | 0.096 |
| *** |  | 0.81 (0.60, 1.09) | 0.159 | 0.73 (0.50, 1.06) | 0.098 |

^a^ Adjusted for Age, BMI, Townsend deprivation index

^b^Additionally adjusted for Smoking status, Coffee intake, Physical activity (MET/h-wk), Education, Tea intake,

Menopause status, Hormone replacement therapy, Parity

*Further adjusted for sleep duration (h/d)

**Further adjusted for insomnia

*** Further adjusted for sleep apnoea

**Table S12**. Associations of sleep traits with reproductive system cancer incidence in the UK Biobank population excluding participants (PCa cohort: 991; ECa cohort: 153 and EOCa cohort: 100) with less than 2 years of follow-up.

|  | **Events** | **Adjusted HR^a^**  **(95% CI)** | **P-value** | **Adjusted HR^b,c^**  **(95% CI)** | **P-value** |
| --- | --- | --- | --- | --- | --- |
| **Prostate cancer** | | | | | |
| **Sleep duration** | |  |  |  |  |
| <7 h | 1,711 | 0.98 (0.93, 1.04) | 0.541 | 1.00 (0.93, 1.06) | 0.920 |
| 7-8 h | 5,288 | Reference |  | Reference |  |
| >8 h | 586 | 0.90 (0.83, 0.99) | 0.021 | 0.88 (0.79, 0.98) | 0.016 |
| Per 1h |  | 1.00 (0.97, 1.02) | 0.679 | 0.98 (0.96, 1.01) | 0.146 |
| **Chronotype** | |  |  |  |  |
| Morning | 4,200 | Reference |  | Reference |  |
| Evening | 2,347 | 1.00 (0.95, 1.05) | 0.924 | 1.01 (0.95, 1.07) | 0.804 |
| **Insomnia** | |  |  |  |  |
| Never/rarely | 2,291 | Reference |  | Reference |  |
| Sometimes | 3,522 | 1.04 (0.98, 1.10) | 0.164 | 1.03 (0.97, 1.10) | 0.316 |
| Usually | 1,898 | 1.04 (0.98, 1.11) | 0.181 | 1.02 (0.95, 1.10) | 0.623 |
| **Snoring** | |  |  |  |  |
| No | 3,702 | Reference |  | Reference |  |
| Yes | 3,531 | 1.05 (1.00, 1.10) | 0.037 | 1.04 (0.98, 1.09) | 0.217 |
| * |  | 1.05 (1.00, 1.10) | 0.033 | 1.04 (0.98, 1.10) | 0.189 |
| ** |  | 1.05 (1.00, 1.10) | 0.038 | 1.03 (0.98, 1.09) | 0.221 |
| *** |  | 1.05 (1.00, 1.10) | 0.033 | 1.04 (0.98, 1.10) | 0.191 |
| **Endometrial cancer** | | | | | |
| **Sleep duration** | |  |  |  |  |
| <7 h | 225 | 0.98 (0.84, 1.14) | 0.769 | 0.89 (0.73, 1.09) | 0.267 |
| 7-8 h | 606 | Reference |  | Reference |  |
| >8 h | 86 | 1.05 (0.84, 1.32) | 0.649 | 0.96 (0.72, 1.29) | 0.809 |
| Per 1h |  | 1.04 (0.98, 1.10) | 0.212 | 1.03 (0.96, 1.11) | 0.370 |
| **Chronotype** | |  |  |  |  |
| Morning | 523 | Reference |  | Reference |  |
| Evening | 323 | 1.02 (0.89, 1.18) | 0.743 | 1.05 (0.89, 1.25) | 0.554 |
| **Insomnia** | |  |  |  |  |
| Never/rarely | 174 | Reference |  | Reference |  |
| Sometimes | 452 | 0.91 (0.76, 1.09) | 0.301 | 1.00 (0.80, 1.24) | 0.992 |
| Usually | 300 | 0.86 (0.71, 1.04) | 0.114 | 0.97 (0.76, 1.23) | 0.791 |
| **Snoring** | |  |  |  |  |
| No | 552 | Reference |  | Reference |  |
| Yes | 288 | 0.97 (0.84, 1.13) | 0.711 | 0.99 (0.83, 1.18) | 0.904 |
| * |  | 0.98 (0.85, 1.14) | 0.823 | 0.99 (0.82, 1.18) | 0.884 |
| ** |  | 0.97 (0.84, 1.12) | 0.705 | 0.99 (0.82, 1.18) | 0.900 |
| *** |  | 0.98 (0.84, 1.13) | 0.740 | 0.99 (0.83, 1.19) | 0.921 |
| **Ovarian cancer** | | | | | |
| **Sleep duration** | |  |  |  |  |
| <7 h | 141 | 1.09 (0.90, 1.32) | 0.386 | 1.16 (0.91, 1.47) | 0.224 |
| 7-8 h | 382 | Reference |  | Reference |  |
| >8 h | 52 | 1.19 (0.89, 1.59) | 0.244 | 1.23 (0.86, 1.75) | 0.259 |
| Per 1h |  | 1.00 (0.93, 1.08) | 0.963 | 0.98 (0.89, 1.07) | 0.649 |
| **Chronotype** | |  |  |  |  |
| Morning | 322 | Reference |  | Reference |  |
| Evening | 199 | 1.10 (0.92, 1.31) | 0.295 | 1.11 (0.89, 1.37) | 0.359 |
| **Insomnia** | |  |  |  |  |
| Never/rarely | 115 | Reference |  | Reference |  |
| Sometimes | 297 | 0.93 (0.75, 1.16) | 0.533 | 1.17 (0.89, 1.55) | 0.266 |
| Usually | 167 | 0.84 (0.66, 1.06) | 0.143 | 1.04 (0.77, 1.42) | 0.792 |
| **Snoring** | |  |  |  |  |
| No | 398 | Reference |  | Reference |  |
| Yes | 124 | 0.77 (0.62, 0.94) | 0.011 | 0.69 (0.53, 0.89) | 0.004 |
| * |  | 0.76 (0.62, 0.94) | 0.011 | 0.69 (0.54, 0.90) | 0.005 |
| ** |  | 0.76 (0.62, 0.93) | 0.009 | 0.68 (0.52, 0.88) | 0.003 |
| *** |  | 0.77 (0.62, 0.94) | 0.011 | 0.69 (0.53, 0.89) | 0.005 |

^a^ Adjusted for Age, BMI, Townsend deprivation index

^b^ Additionally adjusted for Smoking status, Coffee intake, Physical activity (MET/h-wk), Education, Tea intake, History of PSA testing

^c^Additionally adjusted for Smoking status, Coffee intake, Physical activity (MET/h-wk), Education, Tea intake,

Menopause status, Hormone replacement therapy, Parity

*Further adjusted for sleep duration (h/d)

**Further adjusted for insomnia

*** Further adjusted for sleep apnoea

**Table S13**. Associations of sleep traits with reproductive system cancer incidence in the UK Biobank population excluding those with sleep disorders (G47) and jobs involved night shifts and adjusting further for alcohol intake.

|  | **Events** | **Adjusted HR^a^**  **(95% CI)** | **P-value** |
| --- | --- | --- | --- |
| **Prostate cancer** | | | |
| **Sleep duration** | |  |  |
| <7 h | 1,919 | 0.99 (0.93, 1.05) | 0.638 |
| 7-8 h | 5,962 | Reference |  |
| >8 h | 685 | 0.91 (0.82, 1.00) | 0.046 |
| Per 1h |  | 0.99 (0.96, 1.01) | 0.298 |
| **Chronotype** | |  |  |
| Morning | 4,772 | Reference |  |
| Evening | 2,634 | 1.00 (0.95, 1.06) | 0.868 |
| **Chronotype** |  |  |  |
| Definitely morning | 2,037 | Reference |  |
| More morning | 2,735 | 0.99 (0.93, 1.06) | 0.787 |
| More evening | 2,031 | 1.00 (0.93, 1.07) | 0.951 |
| Definitely evening | 603 | 1.01 (0.91, 1.02) | 0.845 |
| Mixed | 1,101 | 1.02 (0.93, 1.11) | 0.673 |
| **Insomnia** | |  |  |
| Never/rarely | 2,441 | Reference |  |
| Sometimes | 3,988 | 1.05 (0.99, 1.11) | 0.101 |
| Usually | 2,164 | 1.03 (0.96, 1.10) | 0.419 |
| No (ref) vs Yes | 6,152 | 1.04 (0.99-1.10) | 0.133 |
| **Daytime sleepiness** |  |  |  |
| Never/rarely | 6,204 | Reference |  |
| Sometimes | 2,098 | 0.99 (0.93, 1.05) | 0.651 |
| Often | 267 | 0.96 (0.83, 1.12) | 0.601 |
| Per category increase | 8,569 | 0.98 (0.94, 1.03) | 0.520 |
| **Daytime napping** |  |  |  |
| Never/rarely | 4,056 | Reference |  |
| Sometimes | 3,819 | 0.97 (0.92, 1.02) | 0.267 |
| Usually | 720 | 0.99 (0.90, 1.09) | 0.844 |
| Per category increase | 8,594 | 0.98 (0.95,1.03) | 0.770 |
| **Snoring** | |  |  |
| No | 4,182 | Reference |  |
| Yes | 3,979 | 1.03 (0.98, 1.09) | 0.197 |
| * |  | 1.04 (0.98, 1.09) | 0.171 |
| ** |  | 1.03 (0.98, 1.09) | 0.194 |
| *** |  | 1.04 (0.98, 1.09) | 0.188 |
| **Endometrial cancer** | | | |
| **Sleep duration** | |  |  |
| <7 h | 258 | 0.88 (0.73, 1.06) | 0.182 |
| 7-8 h | 710 | Reference |  |
| >8 h | 99 | 0.94 (0.71, 1.24) | 0.659 |
| Per 1h |  | 1.05 (0.98, 1.12) | 0.177 |
| **Chronotype** | |  |  |
| Morning | 604 | Reference |  |
| Evening | 384 | 1.06 (0.91, 1.25) | 0.424 |
| **Chronotype** |  |  |  |
| Definitely morning | 264 | Reference |  |
| More morning | 340 | 0.99 (0.81, 1.21) | 0.935 |
| More evening | 290 | 1.09 (0.89, 1.34) | 0.400 |
| Definitely evening | 94 | 0.96 (0.72, 1.32) | 0.846 |
| Mixed | 79 | 0.90 (0.64, 1.26) | 0.549 |
| **Insomnia** | |  |  |
| Never/rarely | 193 | Reference |  |
| Sometimes | 544 | 1.07 (0.87, 1.32) | 0.503 |
| Usually | 341 | 1.02 (0.81, 1.27) | 0.877 |
| No (ref) vs Yes | 885 | 1.08 (0.86, 1.28) | 0.613 |
| **Daytime sleepiness** |  |  |  |
| Never/rarely | 822 | Reference |  |
| Sometimes | 218 | 0.78 (0.64, 0.95) | 0.012 |
| Often | 35 | 0.96 (0.61-1.53) | 0.877 |
| Per category increase | 1,075 | 0.85 (0.72, 1.00) | 0.054 |
| **Daytime napping** |  |  |  |
| Never/rarely | 606 | Reference |  |
| Sometimes | 424 | 0.96 (0.82, 1.12) | 0.607 |
| Usually | 48 | 0.97 (0.66, 1.44) | 0.892 |
| Per category increase | 1,078 | 0.97 (0.85, 1.11) | 0.647 |
| **Snoring** | |  |  |
| No | 642 | Reference |  |
| Yes | 344 | 1.03 (0.87, 1.22) | 0.694 |
| * |  | 1.03 (0.87, 1.22) | 0.719 |
| ** |  | 1.03 (0.87, 1.22) | 0.693 |
| *** |  | 1.03 (0.87, 1.22) | 0.699 |
| **Ovarian Cancer** | | | |
| **Sleep duration** | |  |  |
| <7 h | 165 | 1.16 (0.93, 1.44) | 0.191 |
| 7-8 h | 447 | Reference |  |
| >8 h | 62 | 1.22 (0.88, 1.69) | 0.241 |
| Per 1h |  | 0.97 (0.89, 1.05) | 0.450 |
| **Chronotype** | |  |  |
| Morning | 380 | Reference |  |
| Evening | 228 | 1.07 (0.87, 1.30) | 0.495 |
| **Chronotype** |  |  |  |
| Definitely morning | 162 | Reference |  |
| More morning | 218 | 0.97 (0.76, 1.24) | 0.819 |
| More evening | 165 | 0.98 (0.75, 1.27) | 0.869 |
| Definitely evening | 63 | 1.32 (0.93, 1.87) | 0.118 |
| Mixed | 61 | 0.98 (0.66, 1.46) | 0.925 |
| **Insomnia** | |  |  |
| Never/rarely | 144 | Reference |  |
| Sometimes | 341 | 1.03 (0.80, 1.32) | 0.817 |
| Usually | 194 | 0.94 (0.72, 1.24) | 0.677 |
| No (ref) vs Yes | 535 | 1.00 (0.79, 1.26) | 0.987 |
| **Daytime sleepiness** |  |  |  |
| Never/rarely | 512 | Reference |  |
| Sometimes | 151 | 1.36 (1.10, 1.70) | 0.005 |
| Often | 14 | 0.98 (0.50, 1.90) | 0.945 |
| Per category increase | 677 | 1.22 (1.02, 1.46) | 0.030 |
| **Daytime napping** |  |  |  |
| Never/rarely | 418 | Reference |  |
| Sometimes | 235 | 1.06 (0.87, 1.29) | 0.557 |
| Usually | 27 | 1.05 (0.62, 1.76) | 0.866 |
| Per category increase | 680 | 1.05 (0.89, 1.24) | 0.586 |
| **Snoring** | |  |  |
| No | 457 | Reference |  |
| Yes | 152 | 0.79 (0.63, 0.99) | 0.044 |
| * |  | 0.80 (0.63, 1.00) | 0.054 |
| ** |  | 0.78 (0.62, 0.99) | 0.037 |
| *** |  | 0.79 (0.63, 1.00) | 0.047 |

^a^ Adjusted for age, BMI, Townsend deprivation index, smoking status, coffee intake, physical activity (MET h/wk), education, tea intake, (male-specific) history of PSA testing, (female-specific) menopause status, hormone replacement therapy, and parity.

*Further adjusted for sleep duration (h/d).

**Further adjusted for insomnia.

*** Further adjusted for sleep apnoea.

**Table S14**. Two-sample Mendelian randomisation estimates for the association between sleep traits and risk of reproductive system cancers using sex-combined genetic variants.

| Exposure | | Chronotype | | | Insomnia | | | Snoring | | | Sleep duration | | |
| --- | --- | --- | --- | --- | --- | --- | --- | --- | --- | --- | --- | --- | --- |
| Outcome | **Method** | **SNPs** | **OR (95% CI)** | ***P*** | **SNPs** | **OR (95% CI)** | ***P*** | **SNPs** | **OR (95% CI)** | ***P*** | **SNPs** | **OR (95% CI)** | ***P*** |
| Endometrial cancer | IVW random-effects model | 184 | 0.927 (0.845-1.016) | 0.105 | 61 | 0.921 (0.658-1.290) | 0.633 | 29 | 1.430 (0.577-3.546) | 0.440 | 61 | 0.991 (0.747-1.316) | 0.951 |
|  | Weighted median |  | 0.882 (0.776-1.003) | 0.056 |  | 0.757 (0.465-1.230) | 0.261 |  | 1.078 (0.344-3.378) | 0.898 |  | 0.985 (0.647-1.500) | 0.944 |
|  | Weighted mode |  | 0.833 (0.631-1.100) | 0.200 |  | 0.453 (0.118-1.741) | 0.250 |  | 1.026 (0.141-7.492) | 0.980 |  | 1.057 (0.526-2.126) | 0.876 |
|  | MR-Egger |  | 1.001 (0.786-1.275) | 0.993 |  | 1.043 (0.305-3.566) | 0.947 |  | 0.133 (0.001-14.12) | 0.404 |  | 1.277 (0.439-3.714) | 0.655 |
|  | MR-Egger intercept |  |  | 0.499 |  |  | 0.838 |  |  | 0.318 |  |  | 0.630 |
|  | MR PRESSO |  | 0.919 (0.839-1.006) | 0.070 |  | 0.948 (0.681-1.319) | 0.753 |  | NA |  |  | NA |  |
| Endometrial cancer (endometroid histology) | IVW random-effects model | 207 | 0.969 (0.875-1.074) | 0.553 | 61 | 0.789 (0.527-1.181) | 0.249 | 29 | 1.280 (0.442-3.707) | 0.648 | 61 | 0.968 (0.665-1.409) | 0.864 |
|  | Weighted median |  | 0.864 (0.656-1.138) | 0.300 |  | 0.837 (0.482-1.454) | 0.528 |  | 1.210 (0.306-4.790) | 0.786 |  | 0.808 (0.492-1.328) | 0.400 |
|  | Weighted mode |  | 0.972 (0.849-1.113) | 0.678 |  | 1.467 (0.291-7.401) | 0.643 |  | 1.888 (0.154-23.15) | 0.623 |  | 0.755 (0.306-1.860) | 0.543 |
|  | MR-Egger |  | 0.974 (0.725-1.308) | 0.862 |  | 1.057 (0.242-4.619) | 0.942 |  | 0.303 (0.001-78.31) | 0.677 |  | 1.265 (0.308-5.196) | 0.746 |
|  | MR-Egger intercept |  |  | 0.379 |  |  | 0.687 |  |  | 0.608 |  |  | 0.701 |
|  | MR PRESSO |  | 0.982 (0.888-1.086) | 0.726 |  | NA |  |  | NA |  |  | NA |  |
| Endometrial cancer (non-endometroid histology) | IVW random-effects model | 205 | 0.866 (0.697-1.076) | 0.193 | 61 | 1.953 (0.830-4.598) | 0.125 | 29 | 1.385 (0.118-16.210) | 0.795 | 61 | 1.161 (0.500-2.692) | 0.729 |
|  | Weighted median |  | 0.888 (0.637-1.239) | 0.485 |  | 1.038 (0.284-3.799) | 0.955 |  | 0.915 (0.031-27.271) | 0.959 |  | 0.880 (0.281-2.755) | 0.826 |
|  | Weighted mode |  | 1.056 (0.552-2.018) | 0.870 |  | 0.345 (0.016-7.205) | 0.493 |  | 0.235 (0.001-103.320) | 0.645 |  | 0.740 (0.106-5.148) | 0.762 |
|  | MR-Egger |  | 1.657 (0.917-2.993) | 0.096 |  | 1.260 (0.056-28.279) | 0.884 |  | 0.612 (0.000-153486.206) | 0.939 |  | 2.487 (0.111-55.743) | 0.568 |
|  | MR-Egger intercept |  |  | 0.022 |  |  | 0.774 |  |  | 0.897 |  |  | 0.619 |
|  | MR PRESSO |  |  |  |  | NA |  |  | NA |  |  | NA |  |
| Overall Prostate cancer | IVW random-effects model | 157 | 1.053 (1.011-1.109) | 0.040 | 49 | 1.099 (0.875-1.380) | 0.417 | 29 | 1.931 (0.848-4.394) | 0.117 | 49 | 1.048 (0.865-1.269) | 0.631 |
|  | Weighted median |  | 1.032 (0.963-1.106) | 0.377 |  | 1.036 (0.776-1.383) | 0.813 |  | 1.377 (0.643-2.948) | 0.410 |  | 1.005 (0.800-1.264) | 0.963 |
|  | Weighted mode |  | 1.048 (0.910-1.207) | 0.513 |  | 1.019 (0.535-1.940) | 0.954 |  | 1.008 (0.260-3.901) | 0.991 |  | 1.176 (0.720-1.921) | 0.520 |
|  | MR-Egger |  | 0.936 (0.822-1.066) | 0.321 |  | 1.562 (0.685-3.560) | 0.290 |  | 3.623 (0.062-212.609) | 0.541 |  | 0.786 (0.396-1.560) | 0.494 |
|  | MR-Egger intercept |  |  | 0.056 |  |  | 0.386 |  |  | 0.759 |  |  | 0.396 |
|  | MR PRESSO |  | 1.048 (0.997-1.102) | 0.070 |  | 1.061 (0.854-1.318) | 0.592 |  | 1.295 (0.682-2.456) | 0.436 |  | NA |  |
| Aggressive Prostate cancer | IVW random-effects model | 188 | 1.102 (1.024-1.187) | 0.010 | 63 | 1.059 (0.780-1.439) | 0.713 | 36 | 1.369 (0.573-3.274) | 0.480 | 63 | 1.240 (0.975-1.578) | 0.080 |
|  | Weighted median |  | 1.063 (0.953-1.185) | 0.272 |  | 0.892 (0.579-1.376) | 0.607 |  | 1.096 (0.401-2.998) | 0.858 |  | 1.192 (0.842-1.687) | 0.321 |
|  | Weighted mode |  | 1.022 (0.794-1.316) | 0.867 |  | 0.601 (0.212-1.702) | 0.339 |  | 1.020 (0.131-7.971) | 0.985 |  | 0.777 (0.381-1.586) | 0.491 |
|  | MR-Egger |  | 1.049 (0.865-1.273) | 0.625 |  | 0.668 (0.214-2.084) | 0.488 |  | 9.273 (0.090-950.165) | 0.352 |  | 0.558 (0.222-1.403) | 0.220 |
|  | MR-Egger intercept |  |  | 0.592 |  |  | 0.410 |  |  | 0.415 |  |  | 0.084 |
|  | MR PRESSO |  |  |  |  | 1.058 (0.787-1.423) | 0.706 |  | 1.094 (0.507-2.363) | 0.820 |  | NA |  |
| Overall Epithelial Ovarian cancer | IVW random-effects model | 188 | 0.945 (0.873-1.023) | 0.160 | 63 | 0.837 (0.626-1.121) | 0.232 | 35 | 0.924 (0.255-3.356) | 0.905 | 63 | 1.023 (0.731-1.432) | 0.894 |
|  | Weighted median |  | 0.939 (0.846-1.042) | 0.236 |  | 0.655 (0.432-0.993) | 0.046 |  | 1.347 (0.431-4.210) | 0.609 |  | 0.562 (0.156-2.019) | 0.381 |
|  | Weighted mode |  | 0.915 (0.741-1.129) | 0.407 |  | 0.494 (0.174-1.403) | 0.187 |  | 3.395 (0.258-44.755) | 0.359 |  | 1.081 (0.745-1.566) | 0.683 |
|  | MR-Egger |  | 1.025 (0.832-1.262) | 0.819 |  | 0.922 (0.312-2.728) | 0.884 |  | 13.508 (0.014-13240.336) | 0.464 |  | 1.081 (0.598-1.953) | 0.798 |
|  | MR-Egger intercept |  |  | 0.411 |  |  | 0.884 |  |  | 0.443 |  |  | 0.345 |
|  | MR PRESSO |  | 0.935 (0.866-1.009) | 0.085 |  | NA |  |  | 0.819 (0.322-2.087) | 0.679 |  | 1.136 (0.871-1.482) | 0.349 |
| Serous Epithelial Ovarian cancer | IVW random-effects model | 188 | 0.981 (0.896-1.074) | 0.672 | 63 | 0.805 (0.568-1.140) | 0.221 | 35 | 0.745 (0.178-3.121) | 0.687 | 63 | 0.998 (0.690-1.444) | 0.992 |
|  | Weighted median |  | 0.978 (0.867-1.104) | 0.722 |  | 0.854 (0.521-1.399) | 0.530 |  | 2.048 (0.573-7.320) | 0.270 |  | 1.022 (0.662-1.579) | 0.920 |
|  | Weighted mode |  | 0.968 (0.755-1.241) | 0.799 |  | 0.926 (0.294-2.911) | 0.895 |  | 4.495 (0.364-55.559) | 0.250 |  | 1.443 (0.735-2.833) | 0.291 |
|  | MR-Egger |  | 1.115 (0.878-1.417) | 0.373 |  | 1.083 (0.297-3.953) | 0.904 |  | 23.810 (0.012-48652.029) | 0.421 |  | 0.769 (0.187-3.161) | 0.717 |
|  | MR-Egger intercept |  |  | 0.257 |  |  | 0.641 |  |  | 0.379 |  |  | 0.709 |
|  | MR PRESSO |  | 0.976 (0.896-1.063) | 0.580 |  | 0.774 (0.573-1.063) | 0.116 |  | 0.681 (0.241-1.921) | 0.473 |  | 1.114 (0.824-3.046) | 0.485 |

**Table S14** (continue). Two-sample Mendelian randomisation estimates for the association between sleep traits and risk of reproductive system cancers using sex-combined genetic variants.

| Exposure | | Daytime sleepiness | | |  | Daytime napping |  |
| --- | --- | --- | --- | --- | --- | --- | --- |
| Outcome | **Method** | **SNPs** | **OR (95% CI)** | **P** | **SNPs** | **OR (95% CI)** | **P** |
| Endometrial cancer | IVW random-effects model | 34 | 0.924 (0.413-2.064) | 0.846 | 88 | 1.322 (0.878-1.992) | 0.181 |
|  | Weighted median |  | 0.776 (0.242-2.494) | 0.671 |  | 1.087 (0.606-1.947) | 0.780 |
|  | Weighted mode |  | 0.269 (0.025-2.900) | 0.287 |  | 0.763 (0.182-3.195) | 0.712 |
|  | MR-Egger |  | 0.330 (0.009-12.272) | 0.552 |  | 0.857 (0.211-3.477) | 0.829 |
|  | MR-Egger intercept |  |  | 0.571 |  |  | 0.527 |
|  | MR PRESSO |  | NA |  |  | NA |  |
| Endometrial cancer (endometroid histology) | IVW random-effects model | 34 | 1.180 (0.472-2.947) | 0.723 | 88 | 1.471 (0.937-2.309) | 0.094 |
|  | Weighted median |  | 1.380 (0.394-4.831) | 0.614 |  | 1.473 (0.766-2.833) | 0.245 |
|  | Weighted mode |  | 1.315 (0.108-16.042) | 0.831 |  | 1.357 (0.333-5.529) | 0.672 |
|  | MR-Egger |  | 0.655 (0.011-38.806) | 0.840 |  | 1.104 (0.237-5.131) | 0.900 |
|  | MR-Egger intercept |  |  | 0.774 |  |  | 0.703 |
|  | MR PRESSO |  | NA |  |  | NA |  |
| Endometrial cancer (non-endometroid histology) | IVW random-effects model | 34 | 1.68 (0.167-16.891) | 0.659 | 88 | 1.141 (0.348-3.744) | 0.828 |
|  | Weighted median |  | 6.23 (0.25-155.598) | 0.265 |  | 0.735 (0.130-4.162) | 0.727 |
|  | Weighted mode |  | 28.9 (0.08-10008.4) | 0.267 |  | 0.471 (0.022-10.183) | 0.632 |
|  | MR-Egger |  | 0.034 (0.01-1093.4) | 0.528 |  | 0.013 (0.000-0.660) | 0.033 |
|  | MR-Egger intercept |  |  | 0.456 |  |  | 0.022 |
|  | MR PRESSO |  | NA |  |  | NA |  |
| Overall Prostate cancer | IVW random-effects model | 29 | 0.904 (0.450-1.815) | 0.777 | 78 | 0.907 (0.685-1.201) | 0.495 |
|  | Weighted median |  | 0.713 (0.358-1.417) | 0.334 |  | 0.836 (0.586-1.193) | 0.323 |
|  | Weighted mode |  | 0.794 (0.218-2.888) | 0.728 |  | 0.864 (0.379-1.969) | 0.729 |
|  | MR-Egger |  | 0.346 (0.014-8.256) | 0.517 |  | 0.993 (0.365-2.703) | 0.990 |
|  | MR-Egger intercept |  |  | 0.548 |  |  | 0.853 |
|  | MR PRESSO | 27 | 0.930 (0.500-1.720) | 0.812 |  | NA |  |
| Aggressive Prostate cancer | IVW random-effects model | 34 | 0.544 (0.202-1.468) | 0.229 | 96 | 0.958 (0.637-1.440) | 0.837 |
|  | Weighted median |  | 0.321 (0.104-0.990) | 0.048 |  | 0.808 (0.475-1.376) | 0.432 |
|  | Weighted mode |  | 0.105 (0.007-1.495) | 0.106 |  | 0.598 (0.143-2.500) | 0.483 |
|  | MR-Egger |  | 0.074 (0.001-8.363) | 0.288 |  | 0.841 (0.185-3.813) | 0.823 |
|  | MR-Egger intercept |  |  | 0.404 |  |  | 0.861 |
|  | MR PRESSO | 33 | 0.67 (0.26-1.96) | 0.417 |  | NA |  |
| Overall Epithelial Ovarian cancer | IVW random-effects model | 34 | 0.804 (0.253-2.559) | 0.712 |  | 0.664 (0.434-1.016) | 0.059 |
|  | Weighted median |  | 0.673 (0.243-1.865) | 0.446 |  | 0.768 (0.457-1.290) | 0.318 |
|  | Weighted mode |  | 0.366 (0.061-2.204) | 0.280 |  | 0.817 (0.287-2.331) | 0.707 |
|  | MR-Egger |  | 0.138 (0.01-34.639) | 0.487 |  | 0.154 (0.033-0.714) | 0.019 |
|  | MR-Egger intercept |  |  | 0.527 |  |  | 0.055 |
|  | MR PRESSO | 33 | 1.26 (056-2.84) | 0.568 |  | NA |  |
| Serous Epithelial Ovarian cancer | IVW random-effects model | 34 | 0.725 (0.204-2.571) | 0.618 | 96 | 0.621 (0.388-0.994) | 0.047 |
|  | Weighted median |  | 0.724 (0.221-2.367) | 0.593 |  | 0.663 (0.372-1.184) | 0.165 |
|  | Weighted mode |  | 0.698 (0.098-4.959) | 0.722 |  | 0.649 (0.182-2.315) | 0.507 |
|  | MR-Egger |  | 0.079 (0.01-33.108) | 0.416 |  | 0.184 (0.033-1.022) | 0.056 |
|  | MR-Egger intercept |  |  | 0.467 |  |  | 0.152 |
|  | MR PRESSO | 33 | 1.170 (0.460-2.960) | 0.739 |  | NA |  |

**Table S15**. Two-sample Mendelian randomisation estimates for the association between sleep traits and risk of reproductive system cancers using sex-specific genetic variants.

| Exposure | | Chronotype | | | | Insomnia | | | Snoring | | | | Sleep duration | | |
| --- | --- | --- | --- | --- | --- | --- | --- | --- | --- | --- | --- | --- | --- | --- | --- |
| Outcome | **Method** | **SNPs** | | **OR (95% CI)** | ***P*** | **SNPs** | **OR (95% CI)** | ***P*** | **SNPs** | | **OR (95% CI)** | ***P*** | **SNPs** | **OR (95% CI)** | ***P*** |
| Endometrial cancer | IVW random-effects model | 61 | 1.027 (0.854-1.236) | | 0.774 | 5 | 0.925 (0.569-1.504) | 0.752 | 8 | 0.880 (0.238-3.259) | | 0.848 | 13 | 1.038 (0.687-1.569) | 0.858 |
|  | Weighted median |  | 1.119 (0.891-1.406) | | 0.334 |  | 1.030 (0.675-1.573) | 0.891 |  | 0.712 (0.136-3.723) | | 0.687 |  | 0.907 (0.525-1.565) | 0.725 |
|  | Weighted mode |  | 1.129 (0.684-1.865) | | 0.637 |  | 1.247 (0.668-2.328) | 0.527 |  | 0.707 (0.065-7.731) | | 0.785 |  | 0.875 (0.414-1.850) | 0.733 |
|  | MR-Egger |  | 0.866 (0.523-1.436) | | 0.580 |  | 1.984 (0.644-6.110) | 0.318 |  | 0.379 (0.000-2519241.000) | | 0.908 |  | 0.778 (0.200-3.027) | 0.724 |
|  | MR-Egger intercept |  |  | | 0.480 |  |  | 0.245 |  |  | | 0.919 |  |  | 0.671 |
|  | MR PRESSO |  |  | |  |  | NA |  |  | NA | |  |  | NA |  |
| Endometrial cancer (endometroid histology) | IVW random-effects model | 61 | 1.026 (0.835-1.262) | | 0.807 | 5 | 0.917 (0.609-1.381) | 0.679 | 8 | 1.249 (0.267-5.837) | | 0.778 | 13 | 1.049 (0.645-1.707) | 0.847 |
|  | Weighted median |  | 1.140 (0.868-1.497) | | 0.346 |  | 0.930 (0.577-1.500) | 0.766 |  | 1.317 (0.205-8.450) | | 0.771 |  | 0.784 (0.422-1.457) | 0.442 |
|  | Weighted mode |  | 1.301 (0.769-2.199) | | 0.330 |  | 1.185 (0.546-2.574) | 0.690 |  | 1.060 (0.073-15.384) | | 0.967 |  | 0.693 (0.299-1.606) | 0.409 |
|  | MR-Egger |  | 0.963 (0.546-1.700) | | 0.898 |  | 2.063 (0.765-5.564) | 0.248 |  | 1.573 (0.000-1716162) | | 0.963 |  | 0.554 (0.112-2.752) | 0.486 |
|  | MR-Egger intercept |  |  | | 0.815 |  |  | 0.181 |  |  | | 0.981 |  |  | 0.430 |
|  | MR PRESSO |  |  | |  |  | NA |  |  | NA | |  |  | NA |  |
| Endometrial cancer (non-endometroid histology) | IVW random-effects model | 59 | 1.112 (0.696-1.779) | | 0.656 | 5 | 1.279 (0.395-4.144) | 0.682 | 8 | 0.594 (0.013-26.646) | | 0.788 | 13 | 0.631 (0.194-2.055) | 0.445 |
|  | Weighted median |  | 1.366 (0.712-2.624) | | 0.348 |  | 1.287 (0.359-4.613) | 0.698 |  | 0.230 (0.002-27.660) | | 0.547 |  | 0.774 (0.165-3.618) | 0.744 |
|  | Weighted mode |  | 1.408 (0.368-5.390) | | 0.619 |  | 1.636 (0.315-8.501) | 0.589 |  | 0.218 (0.000-298.367) | | 0.692 |  | 0.692 (0.098-4.902) | 0.719 |
|  | MR-Egger |  | 2.440 (0.639-9.315) | | 0.197 |  | 1.009 (0.029-34.772) | 0.996 |  | 718.664 (0.01- 6.0E+22) | | 0.788 |  | 2.328 (0.049-111.066) | 0.677 |
|  | MR-Egger intercept |  |  | | 0.225 |  |  | 0.896 |  |  | | 0.771 |  |  | 0.501 |
|  | MR PRESSO |  |  | |  |  | NA |  |  | NA | |  |  | NA |  |
| Overall Prostate cancer | IVW random-effects model | 24 | 1.040 (0.881-1.226) | | 0.646 | 3 | 1.328 (0.695-2.538) | 0.390 | 1 | 6.373 (1.068-30.018) | | 0.042 | 2 | 0.687 (0.450-1.050) | 0.083 |
|  | Weighted median |  | 1.062 (0.883-1.279) | | 0.522 |  | 1.062 (0.799-1.412) | 0.677 |  | NA | |  |  | NA |  |
|  | Weighted mode |  | 1.073 (0.799-1.442) | | 0.643 |  | 0.975 (0.750-1.269) | 0.870 |  | NA | |  |  | NA |  |
|  | MR-Egger |  | 1.115 (0.713-1.742) | | 0.638 |  | 0.704 (0.041-12.011) | 0.848 |  | NA | |  |  | NA |  |
|  | MR-Egger intercept |  |  | | 0.744 |  |  | 0.726 |  | NA | |  |  | NA |  |
|  | MR PRESSO |  | 1.093 (0.956-1.250) | | 0.208 |  | NA |  |  | NA | |  |  | NA |  |
| Aggressive Prostate cancer | IVW random-effects model | 28 | 1.165 (0.885-1.532) | | 0.276 | 3 | 1.255 (0.738-2.132) | 0.402 | 1 | 46.409 (2.231-965.095) | | 0.013 | 4 | 0.932 (0.492-1.765) | 0.830 |
|  | Weighted median |  | 1.267 (0.909-1.767) | | 0.163 |  | 1.369 (0.870-2.155) | 0.175 |  | NA | |  |  | 0.774 (0.380-1.577) | 0.481 |
|  | Weighted mode |  | 0.859 (0.423-1.743) | | 0.677 |  | 1.709 (0.734-3.981) | 0.340 |  | NA | |  |  | 0.675 (0.317-1.437) | 0.383 |
|  | MR-Egger |  | 1.729 (0.801-3.730) | | 0.175 |  | 1.348 (0.105-17.391) | 0.857 |  | NA | |  |  | 0.260 (0.040-1.676) | 0.292 |
|  | MR-Egger intercept |  |  | | 0.291 |  |  | 0.963 |  | NA | |  |  |  | 0.292 |
|  | MR PRESSO |  |  | |  |  | NA |  |  | NA | |  |  | NA |  |
| Overall Epithelial Ovarian cancer | IVW random-effects model | 55 | 0.857 (0.730-1.007) | | 0.061 | 5 | 0.959 (0.498-1.848) | 0.901 | 8 | 0.665 (0.128-3.450) | | 0.627 | 13 | 0.723 (0.444-1.176) | 0.191 |
|  | Weighted median |  | 0.830 (0.674-1.023) | | 0.080 |  | 1.321 (0.554-3.149) | 0.531 |  | 1.847 (0.340-10.027) | | 0.477 |  | 0.856 (0.511-1.434) | 0.555 |
|  | Weighted mode |  | 0.882 (0.588-1.324) | | 0.547 |  | 1.370 (0.483-3.886) | 0.586 |  | 3.721 (0.270-51.330) | | 0.359 |  | 1.034 (0.526-2.032) | 0.925 |
|  | MR-Egger |  | 0.905 (0.594-1.378) | | 0.643 |  | 1.683 (0.292-9.688) | 0.601 |  | 0.101 (0.01-10140440) | | 0.865 |  | 1.186 (0.227-6.209) | 0.844 |
|  | MR-Egger intercept |  |  | | 0.787 |  |  | 0.546 |  |  | | 0.889 |  |  | 0.551 |
|  | MR PRESSO |  |  | |  |  | NA |  |  | NA | |  |  | NA |  |
| Serous Epithelial Ovarian cancer | IVW random-effects model | 55 | 0.898 (0.736-1.097) | | 0.292 | 5 | 0.941 (0.441-2.006) | 0.874 | 8 | 1.190 (0.188-7.534) | | 0.853 | 13 | 0.667 (0.363-1.224) | 0.191 |
|  | Weighted median |  | 0.879 (0.689-1.120) | | 0.297 |  | 1.031 (0.391-2.722) | 0.951 |  | 4.544 (0.647-31.918) | | 0.128 |  | 1.059 (0.559-2.007) | 0.861 |
|  | Weighted mode |  | 0.867 (0.551-1.364) | | 0.539 |  | 1.015 (0.330-3.119) | 0.980 |  | 7.327 (0.411-130.566) | | 0.217 |  | 1.205 (0.550-2.643) | 0.650 |
|  | MR-Egger |  | 0.928 (0.550-1.567) | | 0.780 |  | 1.246 (0.165-9.399) | 0.845 |  | 19.080 (0.01-39787870) | | 0.845 |  | 2.213 (0.310-15.775) | 0.445 |
|  | MR-Egger intercept |  |  | | 0.896 |  |  | 0.845 |  |  | | 0.854 |  |  | 0.235 |
|  | MR PRESSO |  | NA | |  |  | NA |  |  | NA | |  |  | NA |  |

**Table S15** (continue). Two-sample Mendelian randomisation estimates for the association between sleep traits and risk of reproductive system cancers using sex-specific genetic variants.

| Exposure | | Daytime napping | | |  | Daytime sleepiness |  |
| --- | --- | --- | --- | --- | --- | --- | --- |
| Outcome | **Method** | **SNPs** | **OR (95% CI)** | **P** | **SNPs** | **OR (95% CI)** | **P** |
| Endometrial cancer | IVW random-effects model | 25 | 1.607 (0.822-3.140) | 0.165 | 9 | 2.374 (0.566-9.961) | 0.237 |
|  | Weighted median |  | 1.301 (0.564-3.003) | 0.537 |  | 1.681 (0.337-8.380) | 0.527 |
|  | Weighted mode |  | 1.439 (0.321-6.452) | 0.639 |  | 1.631 (0.084-31.811) | 0.755 |
|  | MR-Egger |  | 0.652 (0.071-6.030) | 0.710 |  | 0.069 (0.000-157.375) | 0.520 |
|  | MR-Egger intercept |  |  | 0.413 |  |  | 0.392 |
|  | MR PRESSO |  | NA |  |  | NA |  |
| Endometrial cancer (endometroid histology) | IVW random-effects model | 25 | 2.071 (0.930-4.615) | 0.075 | 9 | 2.078 (0.530-8.149) | 0.294 |
|  | Weighted median |  | 1.652 (0.622-4.388) | 0.314 |  | 2.466 (0.400-15.189) | 0.331 |
|  | Weighted mode |  | 1.191 (0.295-4.812) | 0.808 |  | 1.987 (0.097-40.768) | 0.668 |
|  | MR-Egger |  | 1.180 (0.081-17.264) | 0.905 |  | 0.065 (0.000-95.024) | 0.486 |
|  | MR-Egger intercept |  |  | 0.670 |  |  | 0.374 |
|  | MR PRESSO |  | NA |  |  |  |  |
| Endometrial cancer (non-endometroid histology) | IVW random-effects model | 25 | 1.018 (0.156-6.626) | 0.985 | 9 | 19.052 (0.197-1844.788) | 0.207 |
|  | Weighted median |  | 1.335 (0.123-14.435) | 0.812 |  | 2.555 (0.019-336.912) | 0.706 |
|  | Weighted mode |  | 1.208 (0.044-33.126) | 0.912 |  | 0.276 (0.000-202.194) | 0.712 |
|  | MR-Egger |  | 0.026 (0.000-12.158) | 0.256 |  | 0.006 (0.000-579868600.000) | 0.702 |
|  | MR-Egger intercept |  |  | 0.232 |  |  | 0.544 |
|  | MR PRESSO |  | NA |  |  | NA |  |
| Overall Prostate cancer | IVW random-effects model | 14 | 0.653 (0.462-0.924) | 0.016 | 6 | 0.405 (0.147-1.116) | 0.080 |
|  | Weighted median |  | 0.704 (0.437-1.133) | 0.148 |  | 0.574 (0.196-1.677) | 0.310 |
|  | Weighted mode |  | 0.720 (0.327-1.582) | 0.428 |  | 0.670 (0.166-2.704) | 0.598 |
|  | MR-Egger |  | 1.148 (0.189-6.980) | 0.883 |  | 6.302 (0.028-1393.592) | 0.541 |
|  | MR-Egger intercept |  |  | 0.544 |  |  | 0.367 |
|  | MR PRESSO |  | NA |  |  | NA |  |
| Aggressive Prostate cancer | IVW random-effects model | 17 | 0.668 (0.390-1.142) | 0.140 | 2 | 0.110 (0.010-0.830) | 0.012 |
|  | Weighted median |  | 0.813 (0.381-1.734) | 0.592 |  | NA |  |
|  | Weighted mode |  | 1.085 (0.277-4.243) | 0.908 |  | NA |  |
|  | MR-Egger |  | 0.086 (0.004-1.779) | 0.133 |  | NA |  |
|  | MR-Egger intercept |  |  | 0.198 |  | NA |  |
|  | MR PRESSO |  | NA |  |  | NA |  |
| Overall Epithelial Ovarian cancer | IVW random-effects model | 25 | 1.225 (0.665-2.254) | 0.515 | 2 | 0.880 (0.271-2.859) | 0.504 |
|  | Weighted median |  | 1.094 (0.521-2.300) | 0.812 |  | NA |  |
|  | Weighted mode |  | 1.077 (0.323-3.597) | 0.904 |  | NA |  |
|  | MR-Egger |  | 1.179 (0.107-12.992) | 0.894 |  | NA |  |
|  | MR-Egger intercept |  |  | 0.975 |  | NA |  |
|  | MR PRESSO |  | NA |  |  | NA |  |
| Serous Epithelial Ovarian cancer | IVW random-effects model | 25 | 1.232 (0.635-2.392) | 0.537 | 8 | 0.614 (0.169-2.234) | 0.459 |
|  | Weighted median |  | 1.462 (0.599-3.568) | 0.404 |  | 0.557 (0.102-3.028) | 0.498 |
|  | Weighted mode |  | 1.446 (0.271-7.719) | 0.670 |  | 0.465 (0.034-6.310) | 0.583 |
|  | MR-Egger |  | 0.919 (0.068-12.425) | 0.950 |  | 0.017 (0.000-19.891) | 0.300 |
|  | MR-Egger intercept |  |  | 0.821 |  |  | 0.348 |
|  | MR PRESSO |  | NA |  |  | NA |  |
